# Exposome factors predict the association between sleep health and depression

**DOI:** 10.64898/2026.06.26.26356679

**Authors:** Wen Liu, Vincent Küppers, Hanwen Bi, Mostafa Mahdipour, Jianxiao Wu, Fateme Samea, Felix Hoffstaedter, Kathrin Wolf, Charlotte von Gall, Agustin Ibanez, Simon B. Eickhoff, Sarah Genon, Somayeh Maleki Balajoo, Masoud Tahmasian

## Abstract

**Background:** Sleep health and depression are multidimensional constructs, yet their shared determinants remain obscure. We aim to identify the key exposome factors in predicting sleep-related depression (SRD).

**Methods:** We integrated regularized canonical correlation analysis with machine-learning models within a nested cross-validation to identify a shared phenotype (i.e., sleep-related depression (SRD)) and to predict SRD from exposome factors in the UK Biobank (n = 64,781). The best-performing model was validated in an independent subsample at baseline and follow-up (n = 8,139) and in a clinical depression subsample (n = 3,046) to assess generalizability. Subsequently, we conducted complementary analyses to assess the exposome’s role in other latent phenotypes of sleep and depression.

**Findings:** We identified a robust multivariate association between sleep and depression in GP1 (canonical r = 0.42). Exposome factors moderately predicted SRD (r = 0.25; 95% CI [0.25, 0.26]; R² = 0.06; 95% CI [0.06, 0.07]; rMSE = 1.11; 95% CI [1.10, 1.11]). The top predictors were less frequency of confiding in others, less vigorous physical activity, passive smoking exposure, and more sedentary television viewing. Out-of-sample validation of the predictive model showed similar patterns in GP2 at baseline, at follow-up, and in clinical depression subsamples. Similarly, less frequency of confiding in others and greater sedentary television viewing were the main predictors of other depression-related profiles, whereas less walking frequency and less time spent outdoors in winter predicted poor sleep-related profiles.

**Interpretation:** Our generalizable predictive model identifies critical modifiable predictors of the association between sleep health and depression that could serve as potential targets for personalized interventions.

## Introduction

Depression is the leading cause of disability worldwide, ranging from subclinical depressive symptoms in the general population to patients with major depressive disorder (MDD). Depression is inherently multidimensional, characterized by substantial heterogeneity across emotional, cognitive, social, biological, somatic, and sleep domains.^[1,2]^ Poor Sleep health (SH) represents pervasive features and a core domain of depression.^[1,3,4]^ Approximately 78% of patients with MDD report insomnia symptoms.^[5]^ SH is a multifaceted construct encompassing sleep duration, continuity, and quality, which support optimal daytime functioning, mental health, and well-being.^[6,7]^ Individuals with sleep disturbances and insomnia have a greater risk for the onset, recurrence, and exacerbation of MDD.^[8–13]^ Meanwhile, sleep disturbances are also considered a diagnostic criterion for MDD^[2]^, reflecting their bidirectional relationship. Beyond symptom-level associations, converging genetic and neuroimaging evidence further underscores a tight coupling between poor SH and MDD,^[14–17]^ supporting a robust, multidimensional relationship and a shared mechanism underlying the sleep-depression nexus. However, prior studies predominantly relied on group-level comparisons or univariate correlations examining individual sleep characteristics (e.g., insomnia) and overall depression (total scores) in isolation, either in the general population or in patients with MDD.^[18–20]^ These approaches overlook inter-individual variability and fail to capture the complex, multifaceted nature of their associations or their generalizability across populations. A multivariate framework is therefore needed to disentangle these overlapping constructs and identify their shared multidimensional structure. Thus, we introduce *sleep-related depression (SRD)* as a novel construct capturing the specific dimension of depressive symptoms that strongly covaries with SH characteristics at the individual level.

The modifiable factors contributing to their shared variation remain poorly understood, particularly at the individual level across both the general population and MDD samples. The exposome encompassing socioeconomic, lifestyle and environmental factors are modifiable exposures^[21]^ that influence the severity of both depression^[22–24]^ and sleep problems.^[6,25,26]^ Poverty, social isolation, and unhealthy lifestyle, including physical inactivity, malnourishing diet, smoking, and high alcohol consumption, are associated with an increased risk of depressive symptoms and onset of MDD,^[22–24,27]^ whereas healthy lifestyle and social support are protective factors.^[27,28]^ Urbanization-related environmental factors characterized as air pollution, larger street network, and land-use density were positively linked to severe depressive symptoms in the general population.^[29]^ Similarly, higher levels of air pollution and noise, as well as artificial light at night and less closely knit communities, were linked to insomnia symptoms and shorter sleep duration.^[30,31]^ In contrast, exposure to natural environments and green spaces appeared to confer protective effects against depression and was linked to better sleep quality.^[32,33]^ Despite well-documented group-level evidence, existing works have examined the exposome factors in relation to either SH or depression.

Consequently, it remains unclear whether and how these factors shape the shared variance between SH and depression, e.g., by predicting SRD.

This study leveraged large-scale data and multivariate association analyses to derive individual-level SRD scores, representing canonical variates of depression that maximized their correlation with corresponding SH canonical variates — thereby translating shared SH–depression variance into an individual-level metric beyond conventional group-level measures. Machine learning (ML) models were then employed to predict SRD scores from exposome and quantify their relative contributions to the shared SH-depression phenotype in the general population. Model generalizability was evaluated in independent general population subsamples (GP), both cross-sectionally and longitudinally, and in a clinical depression subsample (CD). Using complementary analyses, we examined the differential contributions of these factors across additional latent phenotype profiles, including depression-related sleep (DRS), non-sleep-related depression (NSRD), non-depression-related sleep (NDRS), overall SH, and overall depression. This systematic approach enabled us to distinguish exposome factors associated with the shared SH-depression phenotype from those associated with either SH or depression, providing insights into potentially modifiable population-level factors relevant to the prevention of specific phenotypes.

## Methods

### Participants

Data were obtained from the UK Biobank, a population-based cohort comprising 501,938 participants. The UKB data collection was approved by the Northwest Multicenter Research Ethics Committee, and the present re-analyses of UKB were approved by the ethics committee of Heinrich Heine University Düsseldorf (2018-317-RetroDEuA). To calculate SRD scores, we analyzed four domains in both GP and CD: *i)* seven sleep health (SH) components, namely, sleep duration, chronotype, snoring, insomnia, daytime dozing, daytime napping, and ease of getting up in the morning; *ii)* depression, comprising the Recent Depressive Symptoms scale (RDS-4) and the Neuroticism scale (N-12)^[34]^; *iii)* socioeconomic factors, including social support, working status, and deprivation; *iv)* lifestyle, including physical activity, diet, smoking, alcohol consumption, digital use, and sun exposure; and *v)* environmental factors including air pollution, noise pollution, and greenspace and coastal proximity (Supplemental table 1). The MIND diet was suggested to be negatively associated with depression^[35]^. To reduce dimensionality and address multicollinearity among many diet variables in UKB, diet quality was quantified using a 9-unit modified MIND score^[36]^, which was used as an input feature to predict SRD scores.

Variable selection was based on the availability of UKB data. Participants were included if they had complete data across all domains and were excluded if they had a history of other neuropsychiatric disorders, traumatic brain injury, brain tumors, neurosurgeries, or antipsychotic medication use (Supplemental Table 2 and 3). Sample attrition for variable categories were shown in Supplemental Table 4. Eligible participants were divided into four independent subsamples (Figure 1A). The primary subsample was GP1 (n = 64,781) with cross-sectional data, used to derive the SRD scores, ML predictive models, and complementary analyses. Generalizability of predictive models was assessed in independent subsamples using both baseline (GP2 baseline; n = 8,139) and follow-up (GP2 follow-up; mean follow-up 9.2 years; n = 8,139) assessments to evaluate the long-term roles of baseline exposome factors in predicting follow-up SRD scores. We also assessed multivariate associations and the generalizability of predictive models in CD (n = 3,046). Participants in CD either had a documented ICD-10 diagnosis of current depressive episode (F32) or recurrent depressive disorder (F33) upon assessment.

**Figure 1:**
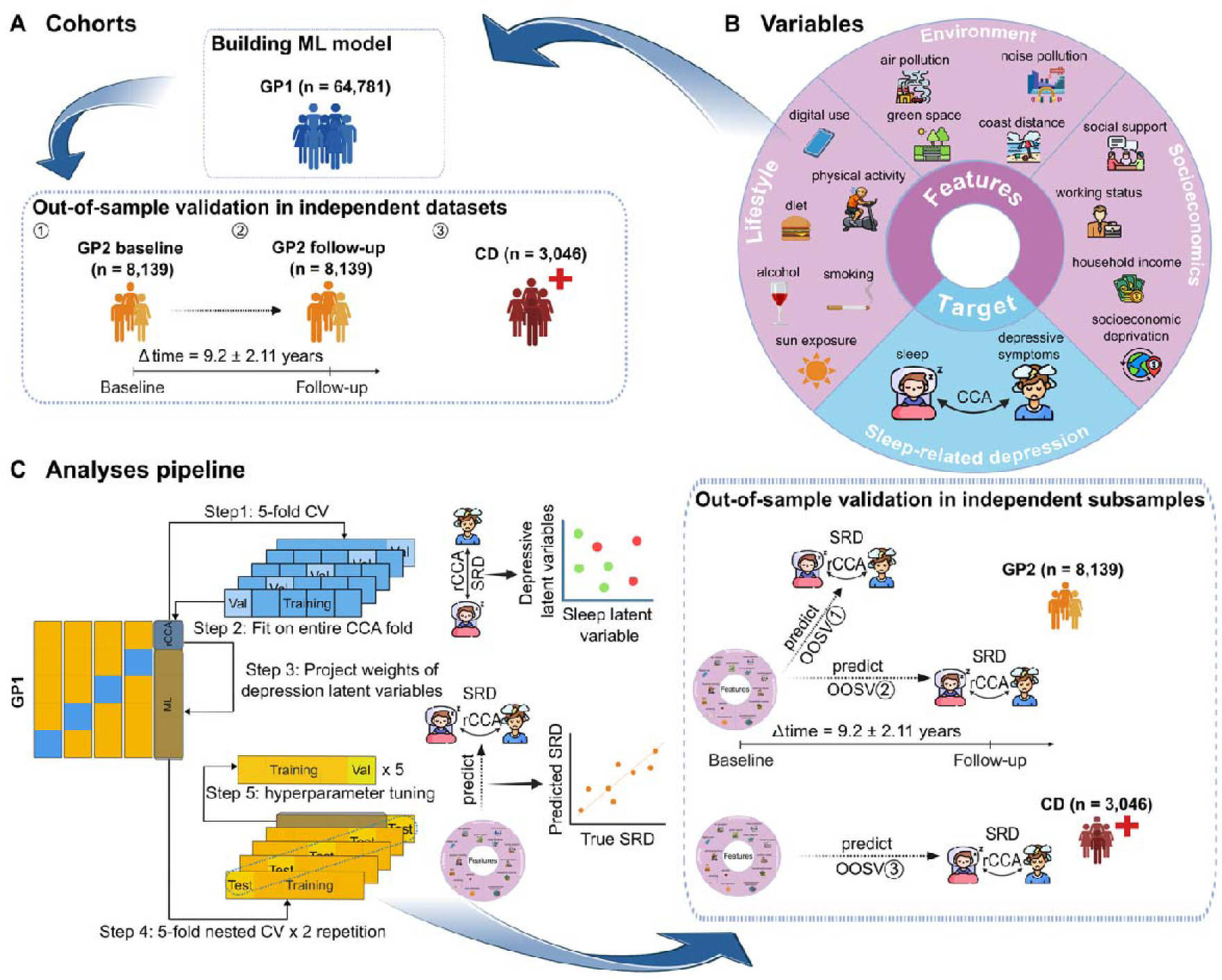
Overview of design and analyses. **A.** The UK Biobank cohort was divided into four independent subsamples. The largest one is for model training and testing (GP1), and three independent subsamples are used for out-of-sample validation (GP2 at baseline and at follow-up, as well as CD). **B.** Overview of variables as exposome factors (input features) and sleep-related depression (SRD) profile (target) in the predictive model. **C.** Regularized canonical correlation analysis (rCCA) identified the strongest multivariate association between SH and depression to obtain the individual-level SRD scores. Subsequently, linear and non-linear machine learning models were trained to predict SRD scores based on exposome factors. The rCCA and predictive procedures were implemented in GP1 within an integrated nested cross-validation framework, with one fold for rCCA and the other four folds for prediction to avoid data leakage. Out-of-sample validation was performed on three independent samples to assess the generalizability of the predictive model trained and tested in GP1. *GP: general population; CD: clinical depression; OOSV: out-of-sample validation; CV: cross-validation; CCA: canonical correlation analysis*.

### Statistical Analyses Analysis framework

To maximize data utilization, obtain unbiased individual-level SRD estimates, and prevent data leakage, we combined regularized canonical correlation analysis (rCCA) and predictive modeling within a nested cross-validation framework^[37]^ (Figure 1C). We randomly split GP1 into five outer folds. In each iteration, one outer fold (CCA fold, 20%) was used to build the rCCA model, while the remaining four folds (ML fold, 80%) were used as training, validation, and test sets for predictive models (Figure 1C). Within each CCA fold, data were further split into a training subset (80%) for hyperparameter tuning and a validation subset (20%) for model selection. In parallel, predictive modeling within each ML fold employed a nested cross-validation scheme^[38]^: We used 80% of the data as an inner training set, and 20% as the outer test set. The inner training set was further subdivided for hyperparameter tuning (80%) and validation (20%). Predictive model performance was evaluated by averaging root mean square error (rMSE) for all ML test sets across all algorithms, and the best-performing algorithm (lowest rMSE) was selected for subsequent analyses. Training and evaluation within ML folds were repeated twice for stability. To prevent data leakage, all preprocessing steps were performed exclusively within the training set. Age and sex were regressed out by fitting linear regression models in the training set, and the resulting regression parameters were applied to the validation and test sets^[39]^. Likewise, continuous and ordinal variables were standardized using the mean and standard deviation estimated from the training set, and these parameters were subsequently applied to the validation and test sets. We applied this procedure consistently to both rCCA and predictive models.

### Regularized Canonical Correlation Analysis

RCCA is a multivariate approach that quantifies the maximum shared variance between at least two sets of variables while reducing overfitting through regularization, as previously applied.^[1,40–42]^ We applied rCCA in GP1 and CD to calculate SRD scores, reflecting the multivariate association between SH and depression characteristics after controlling for age and sex as covariates. The first canonical variate of depression, capturing the dimension most strongly associated with SH, was used to represent individual-level SRD scores. Canonical loadings were calculated as the associations between the original variable and its corresponding canonical variate. Redundancy was calculated as the mean variance of one set explained by the canonical variates of the other set.^[43]^ To minimize overfitting and enhance robustness and generalizability, a regularized version (rCCA) was implemented within a ML framework.^[1,40,44]^ The optimal hyperparameters for rCCA were searched using GridSearchCV (https://scikit-learn.org/stable/modules/generated/sklearn.model_selection.GridSearchCV.html) according to the mean maximized canonical correlation across validation sets.

To assess whether the observed canonical association exceeded the chance level, we evaluated the statistical significance of the multivariate association between sleep health (SH) and depression identified by rCCA using permutation testing. Depression data were independently shuffled across participants for 1,000 iterations, thereby disrupting the original association between SH and depression, while preserving the marginal distributions. For each permutation, the rCCA model with optimised hyperparameters was refit using the shuffled data in the CCA training folds, and the corresponding machine-learning (ML) folds were projected onto the refit rCCA model. Canonical correlation coefficients from both the original and permuted rCCA models were recorded. Empirical *P* values were calculated as the proportion of permuted canonical correlation coefficients exceeding the observed correlation. This procedure was repeated across all ML folds, yielding five *P* values, which were subsequently adjusted for multiple comparisons using false discovery rate (FDR) correction and evaluated at a significance threshold of *P_FDR_* < 0.05.

### Predictive analyses

To evaluate whether exposome factors can predict SRD scores, several linear and non-linear ML models, including ridge regression,^[45]^ LASSO regression,^[46]^ elastic net,^[47]^ random forest,^[48]^ extreme gradient boosting (XGBoost),^[49]^ and linear support vector regression (SVR)^[50]^ were trained in GP1 (Figure 1B). Hyperparameters for predictive models were optimized by the minimal mean error scores across validation sets. Model performance was evaluated by correlation coefficient (r), coefficient of determination (R^2^), and root of mean squared error (rMSE). Statistical significance for the best-performing model was evaluated with a permutation test, where SRD scores were independently shuffled for 500 iterations, thereby disrupting the association between the features and the target. For each permutation, the model with optimized hyperparameters was refit using the shuffled data in the training sets, the test set was then projected onto the refit predictive model. Evaluation metrics for both the original and permuted models were recorded across all test sets. Empirical *P* values were calculated as the proportion of permuted metrics exceeding the observed ones. This procedure was repeated across all test folds, yielding five *P* values, which were subsequently adjusted for multiple comparisons using false discovery rate (FDR) correction and evaluated at a significance threshold of 0.05. To estimate 95% confidence intervals (CI) for the metrics, the participants were randomly resampled for 1000 bootstrap iterations to recalculate the metrics for the CI. Lastly, the best-performing model was calibrated with a linear regression model, in which the real SRD scores were regressed on predicted SRD scores. The calibration slope and intercept were calculated to assess the correspondence between predicted and observed values. To assess whether the predictive model captured target-relevant information beyond demographic characteristics alone, the best-performing model was compared with a demographic model including age, sex, and years of education as predictors. The best-performing model with and without controlling for confounds were compared to evaluate the potential risk from linear confound adjustment into non-linear models^[51]^. Multicollinearity among features was controlled using the variance inflation factor (VIF) before model training, with values > 5 indicating potentially problematic multicollinearity^[52]^.

Feature contributions were quantified using SHapley Additive exPlanations (SHAP) values and examined across age and sex subgroups to assess potential heterogeneity in feature contributions. Given known age- and sex-related differences in both SH and depression^[53,54]^, we assessed whether feature contributions varied by age and sex in the best-performing predictive model. Thus, standardized SHapley Additive exPlanations (SHAP) values were compared between younger (< 65 years) and older (≥ 65 years) participants, as well as between male and female participants. Group differences in SHAP values were evaluated using t tests, with FDR correction for multiple comparisons (P < 0.05). An age cutoff of 65 years was selected based on the statutory retirement age in the United Kingdom^[55]^, as working status was included as a predictor in the model.

### Out-of-sample validation analyses

To evaluate the generalizability of the predictive model across populations, the best-performing predictive model trained in GP1 was then validated in three independent subsamples (GP2 baseline, GP2 follow-up, and CD). For GP2, the depression and SH weights derived from the rCCA analysis in GP1 were applied to the corresponding standardized and de-confounded data in GP2 to project participants onto the latent SH and depression dimensions identified in GP1. Associations between projected SH and depression were then evaluated at GP2 baseline and follow-up to assess the reproducibility and stability of the identified SH–depression relationship in an independent general population cohort. Projection of the GP2 depression data onto the latent dimension identified in GP1 generated SRD scores, reflecting the explicit SH-related depression component. These SRD scores were subsequently used as target variables for the predictive model. As the distribution and multivariate association between SH and depression differed between GP1 and clinical samples, the rCCA model was retrained within CD to obtain a sample-specific SRD phenotype. For out-of-sample validation, the best-performing predictive model trained in GP1 was applied to GP2 (predicting follow-up SRD using baseline predictors) and to subjects in CD using CD-specific SRD scores.

### Complementary analyses

To examine whether and how the same exposome factors contributed differently to predicting other profiles of distinct latent phenotypes, including DRS, NSRD, NDRS, overall depression, and overall SH (Supplementary Methods). The same exposome factors were applied to GP1 using XGBoost to predict each latent profile. To derive latent dimensions comparable to SRD, principal component analysis was performed on each profile, and the first component, representing the largest proportion of variance, was selected as the representative dimension.

We also conducted sensitivity predictive analyses in CD by excluding participants with 1) recurrent depression (n = 77) and 2) both recurrent depression and antidepressant use (n = 1,183, Supplemental Table 5) to control for their impact.

## Results

### Demographics and phenotypic characteristics

The primary rCCA, ML analyses, and complementary analyses were conducted in GP1. Out-of-sample validation was performed in baseline and follow-up GP2 and CD (demographic and feature characteristics in Supplemental 6).

### Multivariate associations between SH and depression

The first canonical dimension captured the strongest shared variance (17.6%) between SH and depression. Our rCCA results in GP1 showed a positive association between the corresponding canonical variates (canonical correlation r_mean_ = 0.42 [SD: 0.002], *P_FDR_* < 0.001; Figure 2A). The canonical variates of depression from this dimension were therefore used to measure SRD scores. These individual-level SRD scores were used as the target for predictive models.

**Figure 2:**
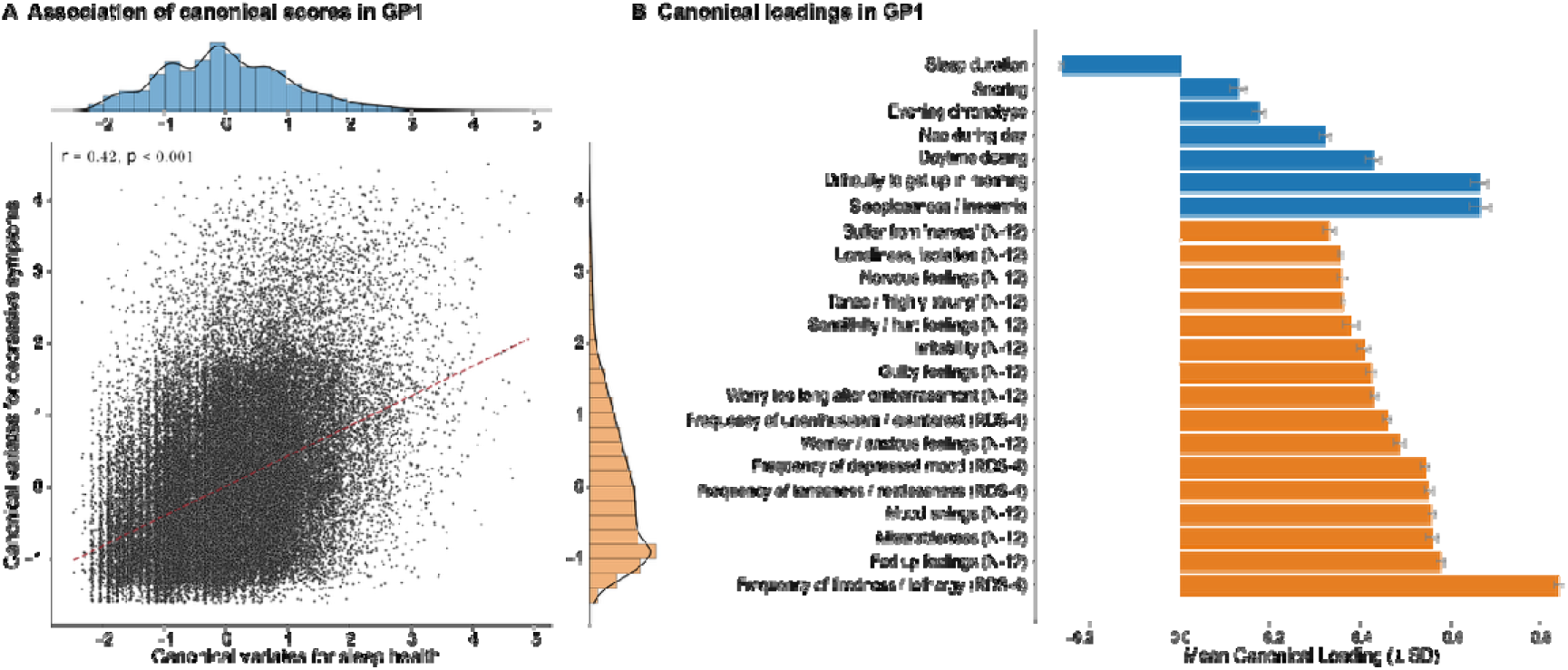
First significant latent dimension and loadings for sleep health (SH) and depression in the general population 1 (GP1) to calculate the sleep-related depression (SRD) profile. A. Multivariate associations between canonical variates for SH and canonical variates of depression in GP1. Each point represents an individual-level SRD score, with scores averaged across the test sets. Model performance is illustrated using the correlation coefficient (r). The blue and orange bar plots show the distributions of latent scores for SH and depression, respectively. B. Mean loadings across five outer CCA folds. Error bars indicate one standard deviation. Greater disturbances in SH loadings were associated with higher depression loadings. The blue bars represent SH, and the orange ones reflect depression. *N-12: neuroticism-12; RDS-4: recent depressive symptoms, indicating depressive symptoms in the past two weeks*.

All SH components were positively associated with depression, except for sleep duration, for which shorter duration was associated with more severe depression. Among all components in SH, insomnia and difficulty getting up in the morning contributed most strongly to the SH variate, whereas frequency of tiredness or lethargy over the past two weeks showed the highest loading among depression measures. Collectively, an individual with a higher SRD score had more sleep disturbances and greater depression severity (Figure 2B). The proportion of SH variance explained by SRD in GP1 was in Supplemental Table 7.

### Predicting SRD from exposome factors in GP1

We assessed multicollinearity among input features prior to ML model training, and all VIF values were < 4, indicating no strong multicollinearity (Supplemental Table 8). The distributions of SRD derived from the five ML folds are shown in Supplemental Figure 1. Across seven ML algorithms to evaluate whether the SRD scores could be predicted from exposome factors in GP1 after controlling age and sex, eXtreme Gradient Boosting (XGBoost) demonstrated the best predictive performance (r: 0.25 ± 0.011; 95% CI: [0.25, 0.26]; *P_FDR_* = 0.002; R²: 0.06 ± 0.006; 95% CI: [0.06, 0.07]; *P_FDR_* = 0.002; rMSE: 1.11 ± 0.014; 95% CI: [1.10, 1.11]; *P_FDR_* = 0.002; Figure 3A, Supplemental Figure 2). XGBoost was calibrated with a slope of 1.05 and an intercept close to 0 (< 0.001, eFigure 3). Feature importance from SHAP analyses revealed that lower social connection, indexed by less frequent confiding in others, was the main predictor of higher SRD scores. This was followed by physical inactivity factors, including fewer days of vigorous physical activity (lasting ≥ 10 minutes/week) and longer hours of sedentary television viewing, and by smoking-related exposure, particularly greater passive smoking outside the home (Figure 3B).

**Figure 3:**
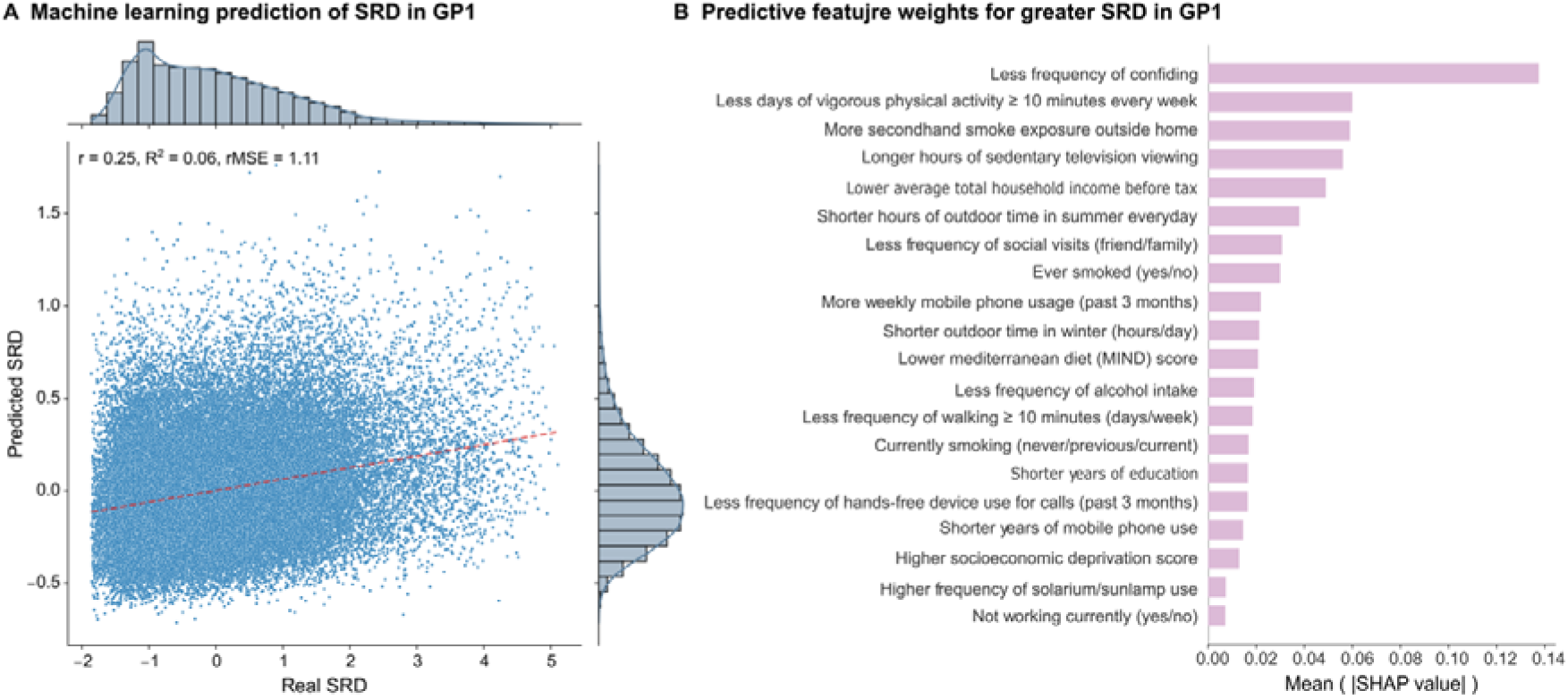
**Predictive model performance in general population 1 (GP1)**. **A.** Each point represents an individual; bar plots represent the distribution of real and predicted SRD scores. Model performance is illustrated using the correlation coefficient (r), coefficient of determination (R²), and root mean square error (rMSE). **B.** Weights for each exposome factors in predicting higher SRD in GP1. The length of each bar represents the mean absolute SHAP values across the split test sets, reflecting the importance of each exposome factor in predicting higher SRD. Only the top 20 features were visualized. *GP: general population subsample; SRD: sleep-related depression; SHAP: SHapley Additive exPlanations*.

Stratified analyses by age and sex demonstrated distinct age- and sex- patterns in the relative importance of exposome factors. Vigorous physical activity was more strongly associated with SRD among younger adults (< 65 years) independent of sex, whereas passive smoking exposure was more important in males, independent of age. However, sedentary television viewing was more prominent among both older adults (≥ 65 years) and females (Supplementary Figure 4 and 5). Feature contribution rankings were largely stable across age and sex subgroups, though FDR-corrected analyses revealed subgroup-specific differences in contribution weights (Supplemental Table 8 and 9). The predictive model outperformed the demographic model (Supplemental Table 10), suggesting that the exposome factors captured meaningful predictive information beyond demographic characteristics alone. Model performances were comparable with and without linear confound adjustment, indicating minimal risk of data leakage (Supplemental Table 11).

### Out-of-sample validation in independent subsamples

Generalizability was evaluated by applying the GP1-trained models to independent datasets. In both GP2 subsamples, the rCCA model trained in GP1 was applied directly to derive the SRD scores, which were subsequently used as the prediction targets. However, in CD, stronger multivariate associations between SH and depression were observed (canonical correlation *r_mean_* = 0.47 [SD, 0.013]; Supplemental Figure 6), indicating a different structure of the SRD phenotype in clinical patients. To avoid underestimating SRD severity in CD and to ensure a representative prediction target, rCCA was retrained within CD using the same framework as GP1, prior to applying the GP1-trained XGBoost model.

Predictive performance was comparable to GP1 in GP2 at baseline for predicting SRD scores, but lower in GP2 for predicting follow-up SRD scores based on baseline features, while with better predictive performance observed in CD (Supplemental Table 12, Figure 4A). Across all validation subsamples, the top predictors identified in GP1 remained largely consistent, with frequency of confiding, frequency of vigorous physical activities, secondhand smoking exposure outside home, and sedentary television viewing ranking among the strongest predictors. (Figure 4B). The proportion of SH explained by SRD in CD was reported in Supplemental Table 12.

**Figure 4:**
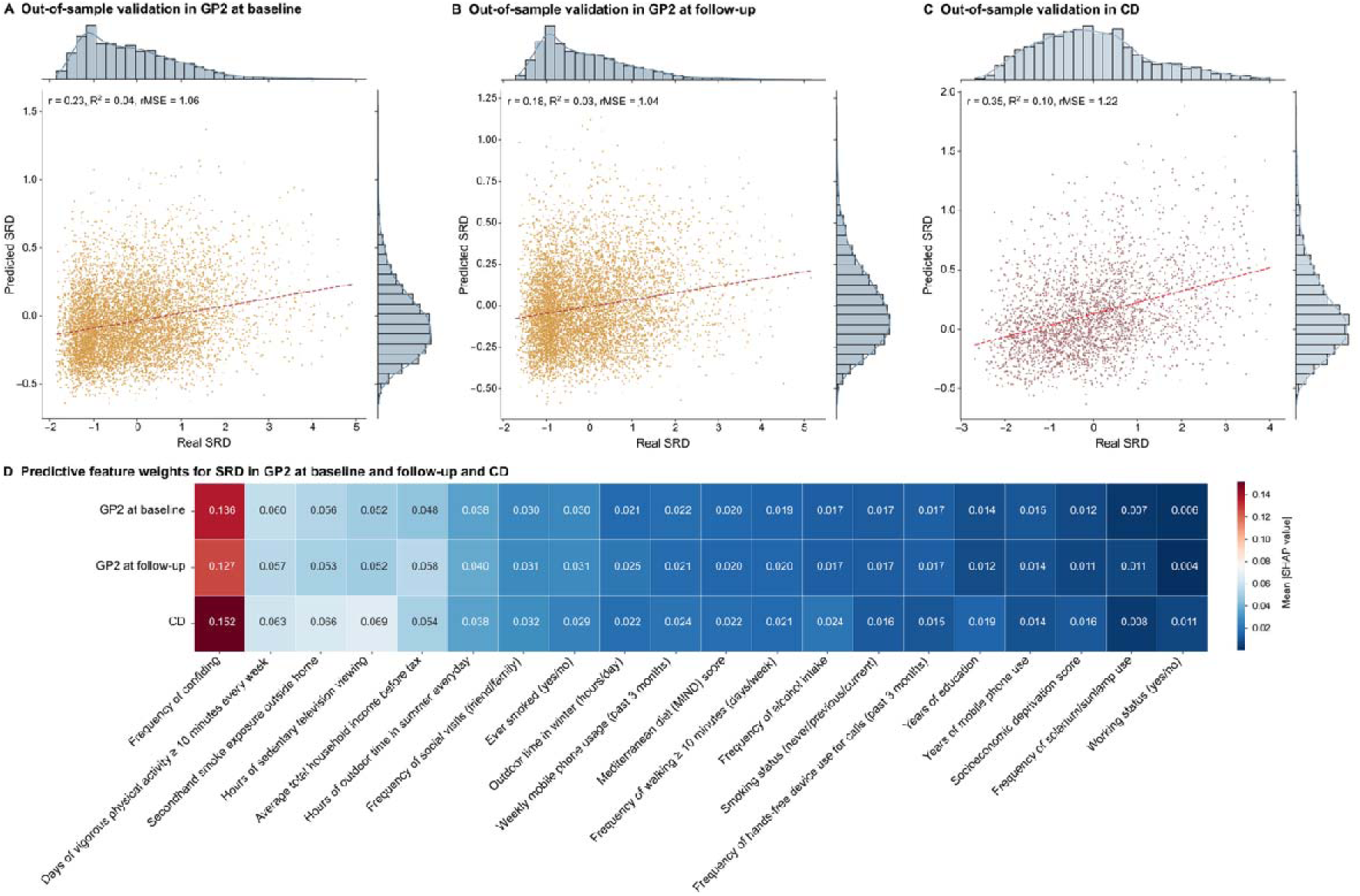
**Out-of-sample validation of the prediction model**. **A and B.** Out-of-sample validation performances in the independent subsamples (GP2) at baseline and follow-up. **C.** Shows out-of- sample validation performance in the CD. Each point represents an individual participant. The bar plots reflect the distribution of real and predicted SRD scores. Model performance is illustrated using the correlation coefficient (r), coefficient of determination (R²), and root mean square error (rMSE). **D.** Weights for each feature in predicting greater SRD in the GP2 at baseline, follow-up, and CD. The colors represent the mean SHAP values for each predictor in the corresponding subsamples. Only the top 20 features were visualized. *GP: general population subsamples; CD: clinical depression subsamples; SRD: sleep-related depression*.

**Figure 5.**
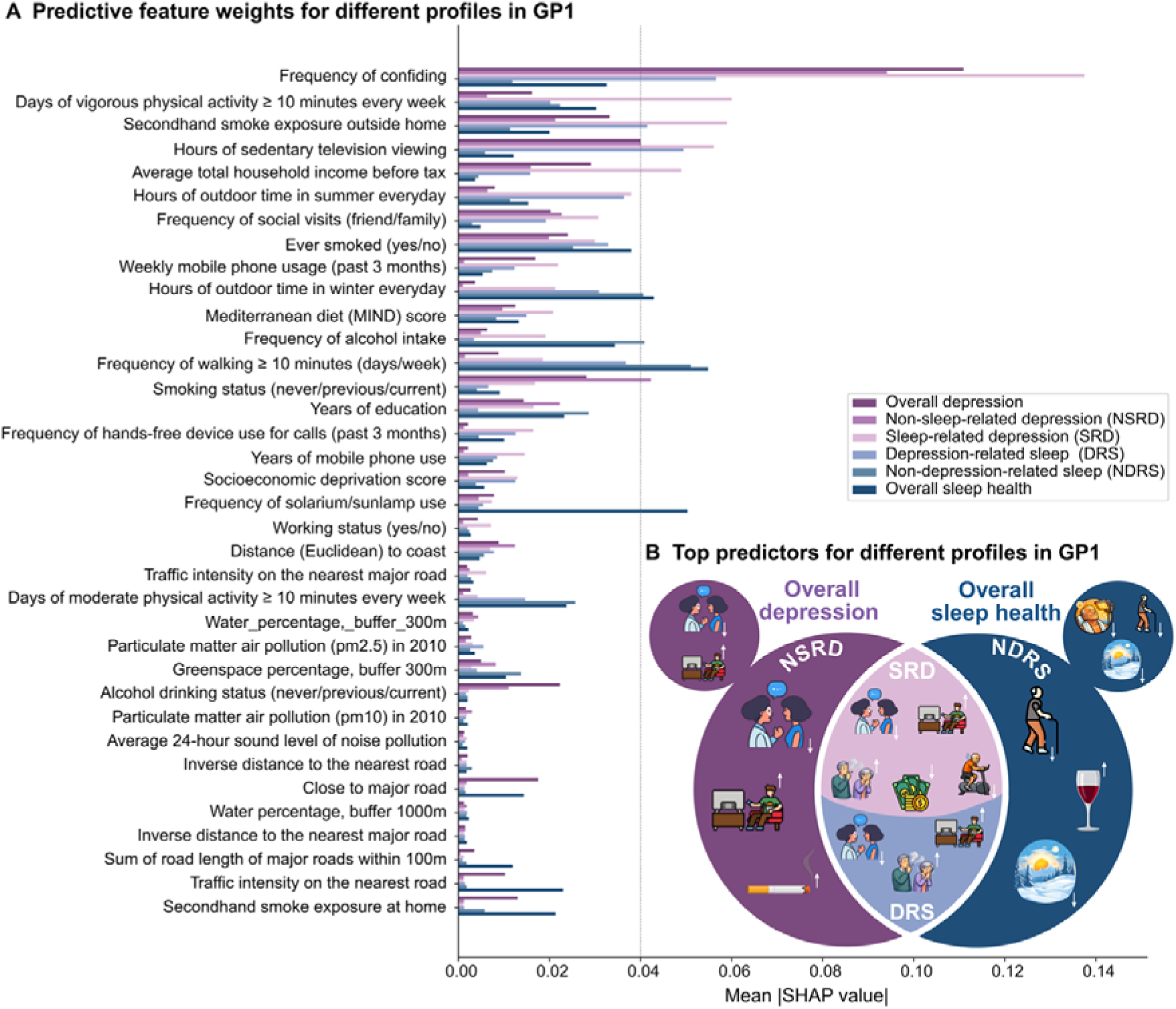
Predictive feature weights for six profiles in GP1. **A.** Feature weights for each exposome factor in predicting different profiles of SH and depression in GP1. Bar length represents the mean absolute SHAP values across the split test sets, reflecting the relative importance of each exposome factor in profile prediction. Colors indicate the different sleep and depression profiles. **B.** This schematic figure illustrates the most important predictors associated with greater symptom severity across six profiles: sleep-related depression (SRD), depression-related sleep (DRS), non-sleep-related depression (NSRD), non-depression-related sleep (NDRS), overall depression (the main dimension of variation in depression), and overall SH (the main dimension of variation in SH). Purple circles represent depression-related profiles, whereas dark blue circles denote sleep-related profiles. *SH: sleep health*.

### Complementary analyses

Lower predictive performance from the same exposome factors was observed in DRS, NDRS, and NSRD, whereas higher performance was identified in overall SH and overall depression (Supplemental Table 13). Similar to SRD, less frequency of confiding and greater sedentary television viewing merged as the primary predictors of *depression-related profiles* (i.e., NSRD and overall depression), whereas less walking frequency, and less time spent outdoors in winter were the main predictors for *poor sleep-related profiles* (i.e., NDRS and overall sleep disturbances). In contrast, more frequent use of artificial light (sunlamps) was a stronger predictor of sleep disturbances, whereas greater frequency of alcohol consumption contributed more strongly to predicting poor NDRS, and current smoking status contributed more strongly to predicting poor NSRD (Figure 5A). These results indicate shared and distinct contributions of exposome factors in predicting SH-specific and depression-specific profiles (Figure 5B).

An identical canonical association between SH and depression was observed in non-recurrent depression (Supplemental Table 14, Supplemental Figure 7), whereas a weaker association was identified in untreated depression (Supplemental Table 14, Supplemental Figure 8). The predictive performance was comparable, and the top predictors remained stable when excluding recurrent depression and excluding recurrent depression and antidepressants (Supplemental Table 15, Supplemental Figures 9 and 10).

## Discussion

In this large-scale study, we employed a novel combined rCCA and ML approach within a nested cross-validation framework to avoid data leakage while maximizing data utilization. We aimed to examine whether a wide range of exposome factors can predict the latent phenotype underlying the interplay between SH and depression. Although the predictive performance of the ML model was moderate, the patterns of contributing exposome factors were broadly consistent and generalizable across cross-sectional and longitudinal GP and CD subsamples. Our rCCA findings identified insomnia and difficulty getting up in the morning as the strongest components linked to depression, such as tiredness and lethargy, in both GP and CD. Despite their robust associations, routine psychotherapies often target either insomnia^[56,57]^ or depression^[1]^ in isolation and fail to alleviate the other condition to achieve full remission, suggesting the importance of simultaneously addressing both conditions. Insomnia is a common residual symptom of depression, which may lead to relapses of depression, suicide, and poor prognosis,^[58,59]^ underscoring the importance of identifying shared influencing factors that may concurrently affect both SH and depression and have investable impacts on treatment outcomes.

Although the protective roles of healthy lifestyle factors such as physical activity, Mediterranean diet, and social support, and the adverse effects of smoking and alcohol on SH and depression are well established,^[23,60–63]^ our results further highlighted their role in predicting SRD. Lower frequency of confiding, fewer days of vigorous physical activity, passive smoking exposure outside the home, and greater hours of sedentary television viewing emerged as the top predictors in the present study.

Consistently, previous studies found that less confiding frequency, less vigorous physical activity (e.g., swimming or cycling), slower walking pace, and longer sedentary television viewing were associated with more depression,^[64,65]^ and confiding frequency together with sedentary television viewing were even suggested as causal factors to depression.^[65]^ Notably, baseline measures of exposome factors moderately predicted subsequent SRD scores assessed approximately 9 years later, underscoring their long-term impacts. The predictive model showed stronger performance in CD, regardless of the effects of recurrent episodes or medication use, suggesting a tighter association between exposome factors and SRD in MDD. This finding was consistent with a meta-analysis reporting larger effect sizes for lifestyle interventions, including physical activity and dietary changes, in MDD patients compared to GP.^[66]^

Although the top predictors identified in the pooled sample remained largely consistent across age- and sex- stratified analyses, several age- and sex-specific patterns emerged. Passive smoking was more prominent in males, whereas vigorous physical activity was more important in younger adults, while sedentary television viewing played a more important role in females and older adults. These findings suggest that age and sex play a non-negligible role in modulating the associations between exposome factors and SRD. Prior studies show that SH and depression profiles drastically vary by age and sex,^[67,68]^ and that physical activities and social support exert different effects across age and sex groups.^[69–71]^ Specifically, walking appears protective against depression in older adults, but not in middle-aged adults^[70]^. Physical activity and instrumental support show greater protective effects in women, whereas emotional support was more important in men.^[69,71]^

Although it has been shown that environmental factors including noise and air pollution are associated with depressive symptoms,^[29]^ they contributed relatively little to predict SRD in our study. This observation is consistent with reports of small effect sizes and limited explained variance (4.71%) for the link between environmental factors and depression.^[29]^ Nevertheless, given the widespread exposure to environmental factors, even relatively small effects may have public health implications and therefore should not be overlooked.

Complementary results also demonstrated distinct predictors for overall depression and overall SH. Socioeconomic and lifestyle factors were more strongly predictive of depression-related profiles than of SH-related profiles. Less frequency of confiding and more sedentary television viewing contributed more strongly to different *depression-related profiles*, whereas less walking frequency and time outdoors in winter showed greater links with *poor SH-related profiles*. Sunlamp use was more strongly related to overall sleep disturbances, while secondhand smoke exposure outside home and sedentary television viewing predicted the shared variance of SH and depression (i.e., SRD and DRS). Current smoking status was more strongly associated with depression after accounting for SH, whereas the frequency of alcohol drinking was more strongly associated with sleep disturbances after accounting for depression. It has been shown that alcohol consumption was significantly related to poor SH, including late chronotype, abnormal duration, snoring, insomnia, and daytime napping.^[72]^ More walking and sunlight exposure have been suggested to be beneficial for reducing sleep disturbances,^[70–74][76,77][73,74][75,76][73,74]^, but less evidence supports an association between exercise intensity and sleep quality improvement.^[75]^ The frequency of confiding in others is internally associated with social isolation and may drive the model in predicting depression-related profiles.^[76]^

### Limitations

These findings should be cautiously interpreted, as the observed effect sizes were modest, accounting for only 6–10% of the variance. The generalizability of our findings based on middle-to-old-aged adults to younger adults and adolescents warrants further investigation. SH was not measured by objective assessment and was not stratified by workdays vs. non-workdays, given the different sleep patterns across these periods.^[26,77,78]^ Moreover, because of the different SH-depression associations in GP and CD, the rCCA model was retrained in CD to derive SRD scores as the target for out-of-sample validation of the predictive model; thus, this is not a fully independent out-of-sample validation in CD. Finally, the current observational design precludes any causal inference.

## Conclusions

Robust multivariate associations between SH and depression were consistently observed across four subsamples, and their shared covariance was moderately predictable from exposome factors.

Maladaptive lifestyle factors, including lower frequency of confiding, greater sedentary television viewing, secondhand smoke exposure, and less vigorous physical activity, emerged as consistent predictors of higher SRD scores across populations and time points. Our systematic approach disentangled the divergent and convergent roles of exposome on the shared and distinct latent dimensions underlying the SH–depression relationships. The generalizability of predictive findings supports the utility of such modifiable factors as potential targets for preventive strategies aimed at reducing the interrelated burden of poor SH and depression.

## Data availability

The data used in this study are derived from the UK Biobank resource were approved by the ethical committee of Heinrich Heine University Düsseldorf (2018-317-RetroDEuA). These data are available under restricted access due to ethical approval requirements, participant consent, and data protection regulations. Access can be obtained by researchers through application to the UK Biobank Access Management System (https://www.ukbiobank.ac.uk/), subject to UK Biobank approval. Raw participant-level data are protected and cannot be publicly shared due to data privacy laws. Processed data that do not contain participant-level information are provided with this paper as Source Data files where applicable. Source data supporting this study are provided with this paper.

## Code availability

The analysis code used in this study is publicly available on GitHub at (https://github.com/wenliu1/Socioeconomic-lifestyle_predicting_sleep_depression_profiles).

## Data Availability

UK Biobank https://www.ukbiobank.ac.uk/

## Acknowledgements

This research was conducted using data from the UK Biobank (https://www.ukbiobank.ac.uk/), a major biomedical database. We are grateful to UK Biobank for providing access to the data and to all study participants for generously contributing their time and information to this resource.

## Author contributions

W.L., S.M.B., and M.T. had full access to all data in the study and took responsibility for the integrity and accuracy of the data and analysis. Study concept and design were by W.L., S.B.E., S.G., A.I., S.M.B., and M.T. Data preparation and preprocessing were carried out by W.L., V.K, F.H. Analyses and model development were conducted by W.L., H.B., J.W., F.S., and S.M.B. Data interpretation was by W.L., S.B.E., S.G., A.I., S.M.B., and M.T. Drafting of the manuscript was by W.L., S.M.B., and M.T. Critical revision of the manuscript for important intellectual content was done by V.K., H.B., J.W., F.S., F.H., C.V.G., S.G., S.B.E, K.W., M.M., and S.G. All authors approved the final version of the manuscript.

## Reference

1. Haritos R, Küppers V, Samea F, Riemann D, Jessen F, Eickhoff SB, et al. The effect of psychotherapy on the multivariate association between insomnia and depressive symptoms in late-life depression. Eur Psychiatry. 2025;68(1):e120. doi:10.1192/j.eurpsy.2025.10088 PubMed PMID: 40898414; PubMed Central PMCID: PMC12438996.

2. Malhi GS, Mann JJ. Depression. The Lancet. 2018;392(10161):2299–312. doi:10.1016/S0140-6736(18)31948-2 PubMed PMID: 30396512.

3. Freeman D, Sheaves B, Waite F, Harvey AG, Harrison PJ. Sleep disturbance and psychiatric disorders. The Lancet Psychiatry. 2020;7(7):628–37. doi:10.1016/S2215-0366(20)30136-X

4. Mendlewicz J. Sleep disturbances: Core symptoms of major depressive disorder rather than associated or comorbid disorders. The World Journal of Biological Psychiatry. 2009;10(4):269–75. doi:10.3109/15622970802503086

5. Toynbee M, Zygmunt D, Levett R, Moghaddacy S, Pappa S. The prevalence of insomnia in depressed adults: a systematic review. Psychiatry Res. 2026;356:116888. doi:10.1016/j.psychres.2025.116888 PubMed PMID: 41389655.

6. Tahmasian M, Küppers V, Genon S, Eickhoff SB, Golombek DA, Ibanez A. Elevating sleep to a global health priority: The One Sleep Health framework. CR Med. 2026;7(6). doi:10.1016/j.xcrm.2026.102828 PubMed PMID: 42173097.

7. Hale L, Troxel W, Buysse DJ. Sleep Health: An Opportunity for Public Health to Address Health Equity. Annu Rev Public Health. 2020;41:81–99. doi:10.1146/annurev-publhealth-040119-094412 PubMed PMID: 31900098; PubMed Central PMCID: PMC7944938.

8. Baglioni C, Battagliese G, Feige B, Spiegelhalder K, Nissen C, Voderholzer U, et al. Insomnia as a predictor of depression: a meta-analytic evaluation of longitudinal epidemiological studies. J Affect Disord. 2011;135(1–3):10–9. doi:10.1016/j.jad.2011.01.011 PubMed PMID: 21300408.

9. Hertenstein E, Benz F, Schneider CL, Baglioni C. Insomnia—A risk factor for mental disorders. Journal of Sleep Research. 2023;32(6):e13930. doi:10.1111/jsr.13930

10. Olfati M, Samea F, Faghihroohi S, Balajoo SM, Küppers V, Genon S, et al. Prediction of depressive symptoms severity based on sleep quality, anxiety, and gray matter volume: a generalizable machine learning approach across three datasets. EBioMedicine. 2024;108:105313. doi:10.1016/j.ebiom.2024.105313 PubMed PMID: 39255547; PubMed Central PMCID: PMC11414575.

11. Tonon AC, Nexha A, Cunningham JEA, d’Eon J, Chakrabarty T, Farzan F, et al. One-Year Actigraphy Study of Sleep and Rest-Activity Rhythms as Markers of Relapse in Depression. JAMA Psychiatry. 2026. doi:10.1001/jamapsychiatry.2025.4453 PubMed PMID: 41670991.

12. Marino C, Andrade B, Montplaisir J, Petit D, Touchette E, Paradis H, et al. Testing Bidirectional, Longitudinal Associations Between Disturbed Sleep and Depressive Symptoms in Children and Adolescents Using Cross-Lagged Models. JAMA Netw Open. 2022;5(8):e2227119. doi:10.1001/jamanetworkopen.2022.27119 PubMed PMID: 35994289; PubMed Central PMCID: PMC9396361.

13. Bao YP, Han Y, Ma J, Wang RJ, Shi L, Wang TY, et al. Cooccurrence and bidirectional prediction of sleep disturbances and depression in older adults: Meta-analysis and systematic review. Neurosci Biobehav Rev. 2017;75:257–73. doi:10.1016/j.neubiorev.2017.01.032 PubMed PMID: 28179129.

14. Sun X, Liu B, Liu S, Wu DJH, Wang J, Qian Y, et al. Sleep disturbance and psychiatric disorders: a bidirectional Mendelian randomisation study. Epidemiology and Psychiatric Sciences. 2022;31:e26. doi:10.1017/S2045796021000810

15. Peng C, Wang K, Wang J, Wassing R, Eickhoff SB, Tahmasian M, et al. Neural correlates of insomnia with depression and anxiety from a neuroimaging perspective: A systematic review. Sleep Med Rev. 2025;81:102093. doi:10.1016/j.smrv.2025.102093 PubMed PMID: 40349510.

16. Gibson MJ, Lawlor DA, Millard LAC. Identifying the potential causal role of insomnia symptoms on 11,409 health-related outcomes: a phenome-wide Mendelian randomisation analysis in UK Biobank. BMC Med. 2023;21(1):128. doi:10.1186/s12916-023-02832-8

17. Wang Y, Tang S, Zhang L, Bu X, Lu L, Li H, et al. Data-driven clustering differentiates subtypes of major depressive disorder with distinct brain connectivity and symptom features. The British Journal of Psychiatry. 2021;219(5):606–13. doi:10.1192/bjp.2021.103

18. Zhu Y, Chen F, Fang X, Liang N, Na G, Liu Z. U-shaped association between sleep duration and depression among postmenopausal women: Evidence from a population-based study. Journal of Affective Disorders. 2026;397:120934. doi:10.1016/j.jad.2025.120934

19. Zhou L, Saltoun K, Carrier J, Storch KF, Dunbar RIM, Bzdok D. Multimodal population study reveals the neurobiological underpinnings of chronotype. Nat Hum Behav. 2025;9(7):1442–56. doi:10.1038/s41562-025-02182-w

20. Lyall LM, Wyse CA, Graham N, Ferguson A, Lyall DM, Cullen B, et al. Association of disrupted circadian rhythmicity with mood disorders, subjective wellbeing, and cognitive function: a cross-sectional study of 91 105 participants from the UK Biobank. Lancet Psychiatry. 2018;5(6):507–14. doi:10.1016/S2215-0366(18)30139-1 PubMed PMID: 29776774.

21. Genon S, Ibanez A, Tahmasian M, Eickhoff SB. Linking the exposome to the brain–behaviour phenotype. Nat Rev Neurosci. 2026;27(7):513–24. doi:10.1038/s41583-026-01049-x

22. van den Bosch M, Meyer-Lindenberg A. Environmental Exposures and Depression: Biological Mechanisms and Epidemiological Evidence. Annual Review of Public Health. 2019;40:239–59. doi:10.1146/annurev-publhealth-040218-044106

23. Firth J, Solmi M, Wootton RE, Vancampfort D, Schuch FB, Hoare E, et al. A meta-review of “lifestyle psychiatry”: the role of exercise, smoking, diet and sleep in the prevention and treatment of mental disorders. World Psychiatry. 2020;19(3):360–80. doi:10.1002/wps.20773

24. Agerbo E, Trabjerg BB, Børglum AD, Schork AJ, Vilhjálmsson BJ, Pedersen CB, et al. Risk of Early-Onset Depression Associated With Polygenic Liability, Parental Psychiatric History, and Socioeconomic Status. JAMA Psychiatry. 2021;78(4):387–97. doi:10.1001/jamapsychiatry.2020.4172

25. Gueye-Ndiaye S, Redline S. Sleep Health Disparities. Annu Rev Med. 2025;76(1):403–15. doi:10.1146/annurev-med-070323-103130 PubMed PMID: 39531860.

26. Aslamyar D, Pilz LK, von Gall C. Relationships Between Self-Reported Sleep Quality, Quantity and Timing on Workdays vs Work-Free Days and Lifestyle Factors in Healthy Adults. Nat Sci Sleep. 2025;17:1641–54. doi:10.2147/NSS.S537593 PubMed PMID: 40688521; PubMed Central PMCID: PMC12276750.

27. Zhao Y, Yang L, Sahakian BJ, Langley C, Zhang W, Kuo K, et al. The brain structure, immunometabolic and genetic mechanisms underlying the association between lifestyle and depression. Nat Mental Health. 2023;1(10):736–50. doi:10.1038/s44220-023-00120-1

28. Zheng YB, Huang YT, Gong YM, Li MZ, Zeng N, Wu SL, et al. Association of lifestyle with sleep health in general population in China: a cross-sectional study. Transl Psychiatry. 2024;14(1):320. doi:10.1038/s41398-024-03002-x

29. Xu J, Liu N, Polemiti E, Garcia-Mondragon L, Tang J, Liu X, et al. Effects of urban living environments on mental health in adults. Nat Med. 2023;29(6):1456–67. doi:10.1038/s41591-023-02365-w

30. Johnson DA, Hirsch JA, Moore KA, Redline S, Diez Roux AV. Associations Between the Built Environment and Objective Measures of Sleep: The Multi-Ethnic Study of Atherosclerosis. Am J Epidemiol. 2018;187(5):941–50. doi:10.1093/aje/kwx302

31. O J, Pugh-Jones C, Clark B, Trott J, Chang L. The Evolutionarily Mismatched Impact of Urbanization on Insomnia Symptoms: a Short Review of the Recent Literature. Curr Psychiatry Rep. 2021;23(5):28. doi:10.1007/s11920-021-01239-7

32. Xie Y, Xiang H, Di N, Mao Z, Hou J, Liu X, et al. Association between residential greenness and sleep quality in Chinese rural population. Environ Int. 2020;145:106100. doi:10.1016/j.envint.2020.106100 PubMed PMID: 32916416.

33. Bratman GN, Hamilton JP, Daily GC. The impacts of nature experience on human cognitive function and mental health. Ann N Y Acad Sci. 2012;1249:118–36. doi:10.1111/j.1749-6632.2011.06400.x PubMed PMID: 22320203.

34. Dutt RK, Hannon K, Easley TO, Griffis JC, Zhang W, Bijsterbosch JD. Mental health in the UK Biobank: A roadmap to self-report measures and neuroimaging correlates. Hum Brain Mapp. 2022;43(2):816–32. doi:10.1002/hbm.25690 PubMed PMID: 34708477; PubMed Central PMCID: PMC8720192.

35. Cherian L, Wang Y, Holland T, Agarwal P, Aggarwal N, Morris MC. DASH and Mediterranean-Dash Intervention for Neurodegenerative Delay (MIND) Diets Are Associated With Fewer Depressive Symptoms Over Time. J Gerontol A Biol Sci Med Sci. 2021;76(1):151–6. doi:10.1093/gerona/glaa044 PubMed PMID: 32080745; PubMed Central PMCID: PMC7973257.

36. Morris MC, Tangney CC, Wang Y, Sacks FM, Barnes LL, Bennett DA, et al. MIND diet slows cognitive decline with aging. Alzheimers Dement. 2015;11(9):1015–22. doi:10.1016/j.jalz.2015.04.011 PubMed PMID: 26086182; PubMed Central PMCID: PMC4581900.

37. Domingos P. A few useful things to know about machine learning. Commun ACM. 2012;55(10):78–87. doi:10.1145/2347736.2347755

38. Berrar D. Cross-Validation. Encyclopedia of Bioinformatics and Computational Biology (Second Edition). 2025;638–44. doi:10.1016/B978-0-323-95502-7.00032-4

39. Komeyer V, Nieto N, Eickhoff SB, Raimondo F, Patil KR. Overview of Challenges in Brain-Based Predictive Modeling: Toward Meaningful Predictive Insights. Biological Psychiatry. 2025. doi:10.1016/j.biopsych.2025.09.003

40. Küppers V, Bi H, Nicolaisen-Sobesky E, Hoffstaedter F, Yeo BTT, Drzezga A, et al. Multivariate associations of motor performance, sleep quality, depressive symptoms, and grey matter volume in younger and mid-to-older adults. Sci Rep. 2026;16(1):1318. doi:10.1038/s41598-025-34951-y

41. Balajoo SM, Plachti A, Nicolaisen-Sobesky E, Dong D, Hoffstaedter F, Meuth SG, et al. Discovery, Replicability, and Generalizability of a Left Anterior Hippocampus’ Morphological Network Linked to Self-Regulation. Hum Brain Mapp. 2024;45(18):e70099. doi:10.1002/hbm.70099 PubMed PMID: 39705057; PubMed Central PMCID: PMC11661003.

42. Nicolaisen-Sobesky E, Maleki Balajoo S, Mahdipour M, Mihalik A, Olfati M, Hoffstaedter F, et al. Cardiometabolic health and physical robustness map onto distinct patterns of brain structure and neurotransmitter systems. PLoS Biol. 2025;23(11):e3003498. doi:10.1371/journal.pbio.3003498 PubMed PMID: 41270115; PubMed Central PMCID: PMC12671889.

43. Stewart D, Love W. A general canonical correlation index. Psychological Bulletin. 1968;70(3, Pt.1):160–3. doi:10.1037/h0026143

44. Mihalik A, Ferreira FS, Moutoussis M, Ziegler G, Adams RA, Rosa MJ, et al. Multiple Holdouts With Stability: Improving the Generalizability of Machine Learning Analyses of Brain–Behavior Relationships. Biological Psychiatry. 2020;Innovations in Clinical Neuroscience: Tools, Techniques, and Transformative Frameworks 87(4):368–76. doi:10.1016/j.biopsych.2019.12.001

45. McDonald GC. Ridge regression. WIREs Computational Statistics. 2009;1(1):93–100. doi:10.1002/wics.14

46. Ranstam J, Cook JA. LASSO regression. Br J Surg. 2018;105(10):1348. doi:10.1002/bjs.10895

47. Zou H, Hastie T. Regularization and Variable Selection Via the Elastic Net. J R Stat Soc Ser B Stat Methodol. 2005;67(2):301–20. doi:10.1111/j.1467-9868.2005.00503.x

48. Breiman L. Random Forests. Machine Learning. 2001;45(1):5–32. doi:10.1023/A:1010933404324

49. Chen T, Guestrin C. XGBoost: A Scalable Tree Boosting System. In: Proceedings of the 22nd ACM SIGKDD International Conference on Knowledge Discovery and Data Mining [Internet]. New York, NY, USA: Association for Computing Machinery; 2016 [cited 2026 Jan 4]. p. 785–94. (KDD ’16). Available from: 10.1145/2939672.2939785 doi: 10.1145/2939672.2939785

50. Awad M, Khanna R. Support Vector Regression. In: Awad M, Khanna R, editors. Efficient Learning Machines: Theories, Concepts, and Applications for Engineers and System Designers [Internet]. Berkeley, CA: Apress; 2015 [cited 2026 Jan 4]. p. 67–80. Available from: 10.1007/978-1-4302-5990-9_4 doi: 10.1007/978-1-4302-5990-9_4

51. Hamdan S, Love BC, von Polier GG, Weis S, Schwender H, Eickhoff SB, et al. Confound-leakage: confound removal in machine learning leads to leakage. Gigascience. 2022;12:giad071. doi:10.1093/gigascience/giad071 PubMed PMID: 37776368; PubMed Central PMCID: PMC10541796.

52. Shrestha N. Detecting Multicollinearity in Regression Analysis. American Journal of Applied Mathematics and Statistics. 2020;8(2):39–42. doi:10.12691/ajams-8-2-1

53. Epidemiology of sleep health. In: Sleep and Health [Internet]. Academic Press; 2026 [cited 2026 Jan 23]. p. 13–22. Available from: https://www.sciencedirect.com/science/chapter/edited-volume/abs/pii/B978044313954300019X doi:10.1016/B978-0-443-13954-3.00019-X

54. Luppa M, Sikorski C, Luck T, Ehreke L, Konnopka A, Wiese B, et al. Age- and gender-specific prevalence of depression in latest-life--systematic review and meta-analysis. J Affect Disord. 2012;136(3):212–21. doi:10.1016/j.jad.2010.11.033 PubMed PMID: 21194754.

55. GOV.UK [Internet]. [cited 2026 Jan 7]. Check your State Pension age. Available from: https://www.gov.uk/state-pension-age

56. Blanken TF, Van Der Zweerde T, Van Straten A, Van Someren EJW, Borsboom D, Lancee J. Introducing Network Intervention Analysis to Investigate Sequential, Symptom-Specific Treatment Effects: A Demonstration in Co-Occurring Insomnia and Depression. Psychother Psychosom. 2019;88(1):52–4. doi:10.1159/000495045

57. Blom K, Forsell E, Hellberg M, Svanborg C, Jernelöv S, Kaldo V. Psychological Treatment of Comorbid Insomnia and Depression: A Double-Blind Randomized Placebo-Controlled Trial. Psychother Psychosom. 2024;93(2):100–13. doi:10.1159/000536063

58. Bei B, Asarnow LD, Krystal A, Edinger JD, Buysse DJ, Manber R. Treating insomnia in depression: Insomnia related factors predict long-term depression trajectories. J Consult Clin Psychol. 2018;86(3):282– 93. doi:10.1037/ccp0000282 PubMed PMID: 29504795; PubMed Central PMCID: PMC5841584.

59. Inada K, Enomoto M, Yamato K, Marumoto T, Takeshima M, Mishima K. Effect of residual insomnia and use of hypnotics on relapse of depression: a retrospective cohort study using a health insurance claims database. Journal of Affective Disorders. 2021;281:539–46. doi:10.1016/j.jad.2020.12.040

60. Grigoriou I, Kotoulas SC, Porpodis K, Spyratos D, Papagiouvanni I, Tsantos A, et al. The Interactions between Smoking and Sleep. Biomedicines. 2024;12(8). doi:10.3390/biomedicines12081765

61. Cruz J, Llodio I, Iturricastillo A, Yanci J, Sánchez-Díaz S, Romaratezabala E. Association of Physical Activity and/or Diet with Sleep Quality and Duration in Adolescents: A Scoping Review. Nutrients. 2024;16(19). doi:10.3390/nu16193345

62. Huang HH, Stubbs B, Chen LJ, Ku PW, Hsu TY, Lin CW, et al. The effect of physical activity on sleep disturbance in various populations: a scoping review of randomized clinical trials. Int J Behav Nutr Phys Act. 2023;20(1):44. doi:10.1186/s12966-023-01449-7

63. Cleary JL, Fang Y, Zahodne LB, Bohnert ASB, Burmeister M, Sen S. Polygenic Risk and Social Support in Predicting Depression Under Stress. Am J Psychiatry. 2023;180(2):139–45. doi:10.1176/appi.ajp.21111100 PubMed PMID: 36628515; PubMed Central PMCID: PMC10355168.

64. Santos J, Ihle A, Peralta M, Domingos C, Gouveia ÉR, Ferrari G, et al. Associations of Physical Activity and Television Viewing With Depressive Symptoms of the European Adults. Front Public Health. 2021;9:799870. doi:10.3389/fpubh.2021.799870 PubMed PMID: 35096747; PubMed Central PMCID: PMC8790035.

65. Choi KW, Stein MB, Nishimi KM, Ge T, Coleman JRI, Chen CY, et al. An Exposure-Wide and Mendelian Randomization Approach to Identifying Modifiable Factors for the Prevention of Depression. Am J Psychiatry. 2020;177(10):944–54. doi:10.1176/appi.ajp.2020.19111158 PubMed PMID: 32791893; PubMed Central PMCID: PMC9361193.

66. Amiri S, Mahmood N, Javaid SF, Khan MA. The Effect of Lifestyle Interventions on Anxiety, Depression and Stress: A Systematic Review and Meta-Analysis of Randomized Clinical Trials. Healthcare. 2024;12(22):2263. doi:10.3390/healthcare12222263

67. Grandner M. Epidemiology of sleep health. In: Sleep and Health [Internet]. Academic Press; 2026 [cited 2026 Feb 4]. p. 13–22. Available from: https://www.sciencedirect.com/science/chapter/edited-volume/abs/pii/B978044313954300019X doi:10.1016/B978-0-443-13954-3.00019-X

68. Negeri ZF, Levis B, Sun Y, He C, Krishnan A, Wu Y, et al. Accuracy of the Patient Health Questionnaire-9 for screening to detect major depression: updated systematic review and individual participant data meta-analysis. BMJ. 2021;375:n2183. doi:10.1136/bmj.n2183 PubMed PMID: 34610915; PubMed Central PMCID: PMC8491108.

69. Zhang J, Yen ST. Physical Activity, Gender Difference, and Depressive Symptoms. Health Serv Res. 2015;50(5):1550–73. doi:10.1111/1475-6773.12285 PubMed PMID: 25630931; PubMed Central PMCID: PMC4600361.

70. Fukukawa Y, Nakashima C, Tsuboi S, Kozakai R, Doyo W, Niino N, et al. Age differences in the effect of physical activity on depressive symptoms. Psychol Aging. 2004;19(2):346–51. doi:10.1037/0882-7974.19.2.346 PubMed PMID: 15222828.

71. Amaike M, Yokoyama A, Tanaka Y, Yamazaki M, Matsuzawa A, Ojima T, et al. Incidence of depressive symptoms and their associations with lifestyle and social support networks among community-dwelling older adults: a sex-stratified longitudinal study using the JAGES study. BioPsychoSocial Med. 2025;19(1):20. doi:10.1186/s13030-025-00342-y

72. Zheng JW, Ai SZ, Chang SH, Meng SQ, Shi L, Deng JH, et al. Association between alcohol consumption and sleep traits: observational and mendelian randomization studies in the UK biobank. Mol Psychiatry. 2024;29(3):838–46. doi:10.1038/s41380-023-02375-7

73. Siraji MA, Spitschan M, Kalavally V, Haque S. Light exposure behaviors predict mood, memory and sleep quality. Sci Rep. 2023;13(1):12425. doi:10.1038/s41598-023-39636-y

74. Wang Y, Yao Z, Wang M, Liu H, Shi S, Zheng F, et al. Daily steps and sleep in adults: A systematic review and meta-analysis. Sleep Medicine. 2026;138:108697. doi:10.1016/j.sleep.2025.108697

75. Liang Z, Zhao S, Tian S, Wang C, Zhang M, Xu K, et al. Determinants and interrelations of exercise effects on sleep quality: A multimethod meta-analysis. Sleep Medicine Reviews. 2026;86:102239. doi:10.1016/j.smrv.2026.102239

76. Liu L, Gou Z, Zuo J. Social support mediates loneliness and depression in elderly people. J Health Psychol. 2016;21(5):750–8. doi:10.1177/1359105314536941 PubMed PMID: 24925547.

77. Gall C von, Holub L, Pfeffer M, Eickhoff S. Chronotype-Dependent Sleep Loss Is Associated with a Lower Amplitude in Circadian Rhythm and a Higher Fragmentation of REM Sleep in Young Healthy Adults. Brain Sci. 2023;13(10):1482. doi:10.3390/brainsci13101482 PubMed PMID: 37891848; PubMed Central PMCID: PMC10605513.

78. Aslamyar D, Gall C von. Sleep Compensation on Work-Free days Is Associated with Better Sleep Quality. Nat Sci Sleep. 2025;17:3137–48. doi:10.2147/NSS.S562192 PubMed PMID: 41426203; PubMed Central PMCID: PMC12717801.

