## Supplement for "Exposome factors predict the association between sleep health and depression"

Liu W. et al.

**Supplementary file**

**Supplemental Methods:**

**Complementary analyses**

To examine how the same exposome factors contribute to other sleep health and depression profiles, we conducted complementary analyses for the following latent profiles, as follows:

- **Depression-related sleep (DRS):** We defined Depression-related sleep (DRS) as a new concept describing the specific dimension of SH that strongly covaries with depression at the individual level. The DRS scores represent the first canonical variate of SH, defined as the linear combination that maximizes its correlation with the corresponding canonical variate of depression at the individual level. We derived DRS from the same rCCA model used to define SRD in the GP1 subsample and then predicted it from the same set of exposome factors using the XGBoost model used to predict SRD.
- **Non-sleep-related depression (NSRD):** We used multivariate generalized linear models and multivariate logistic regression to regress out sleep health, along with age and sex as confounders, from depression variables after standardizing ordinal and categorical variables, respectively. Residualized depression measures were subsequently entered into principal component analysis (PCA) with varimax rotation to identify the first latent dimension independent of SH. The first component, representing the largest proportion of variance in residualized depression, was defined as NSRD and used as the target for the downstream predictive models. To prevent data leakage, we performed all preprocessing and model-fitting procedures within a 5-fold nested cross-validation framework. Our dataset was first divided into 5 outer folds, with each fold iteratively serving as the test set. The remaining 4 outer folds constituted the training set and were further split into inner training (80%) and validation (20%) sets for hyperparameter tuning. Multivariate generalized linear models, logistic regression models, and PCA were trained in the same training set and subsequently applied to the held-out test set. We projected residualized depression measures from the test set onto the PCA model to obtain the first principal component score, which served as the proxy for NSRD. Lastly, we used the best-performing SRD model to predict the first component of residualized depression in the test set, using the same exposome factors and settings as for SRD. Both the predictive model and PCA were integrated into a nested cross-validation framework. Age and sex were regressed out as confounds in both PCA and predictive models.
- **Non-depression-related sleep (NDRS):** We applied the same analytical framework to derive depression-independent SH as a proxy for NDRS, which we then used as the target for downstream predictive modeling.
- **Overall SH:** To obtain the comparable latent dimension of the overall SH for predictive modeling, PCA with varimax rotation was performed within a 5-fold nested cross-validation framework. We then used the first component, which explains the largest proportion of variance in SH scores, as the target for a predictive model based on exposome factors. The PCA and predictive model were embedded into a nested cross-validation framework as described previously to avoid data leakage. Age and sex were regressed out as confounds in both PCA and predictive models.
- **Overall depression:** The same analytical framework was used to derive the first component of depression as described for overall SH.

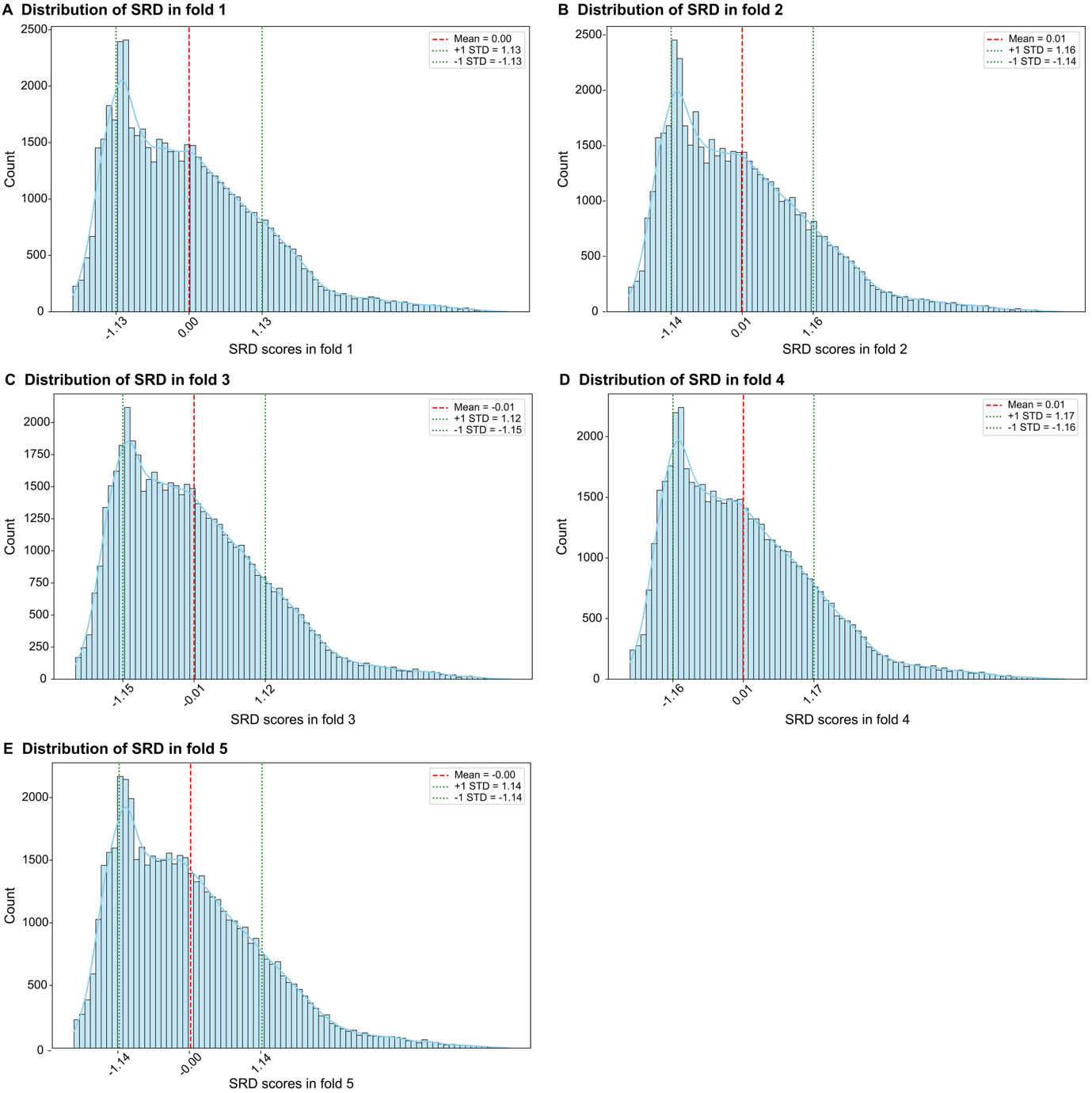

**Supplemental Figure 1. Sleep-related depression (SRD) distribution across five folds.** A-E panels show the distributions of SRD scores across the five folds split in the canonical correlation analysis. Histograms represent the frequency distribution of SRD scores within each fold. Vertical dashed lines indicate the mean ± 1SD of SRD scores for each fold. *SD: standard deviation.*

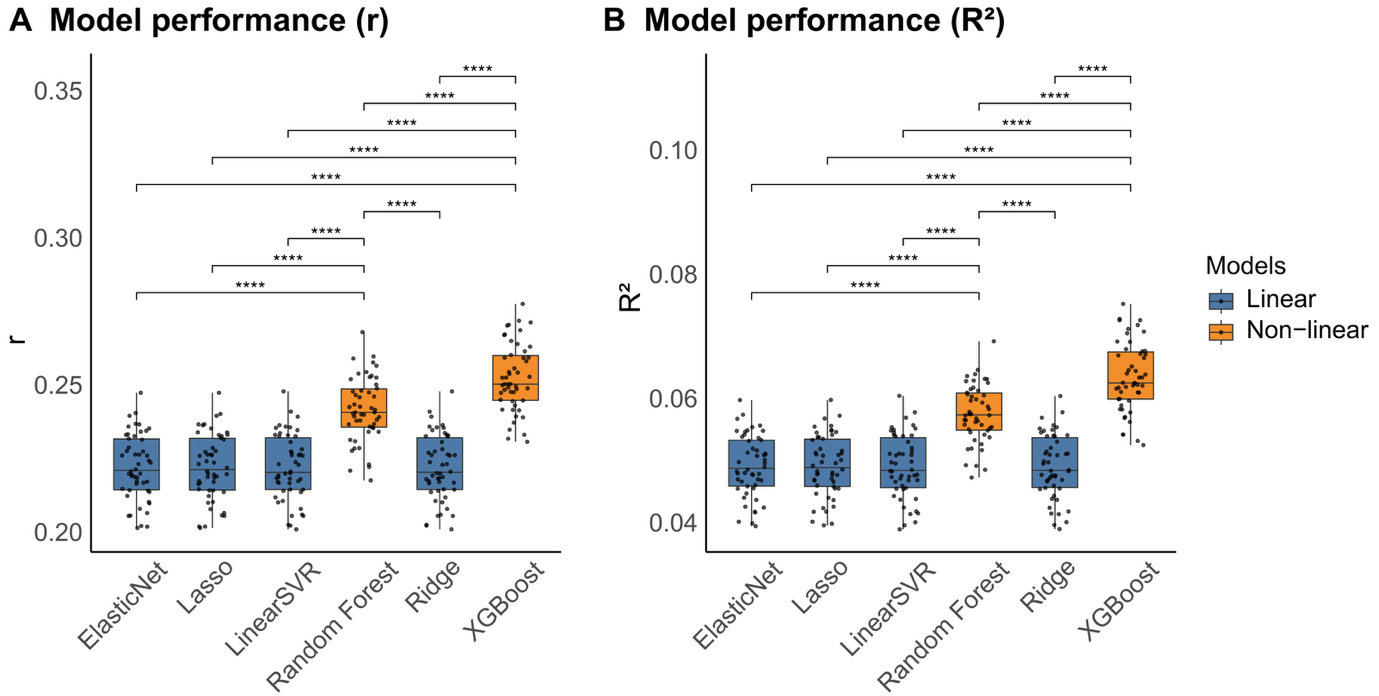

**Supplemental Figure 2. Performance of different linear and non-linear ML models in predicting sleep-related depression (SRD) in the general population 1.** Model performance is illustrated using the correlation coefficient (r; A) and the coefficient of determination (R²; B). Each point represents the metric value (r, R2) for an algorithm. The blue and orange bars represent linear models and non-linear models, respectively. ****: *P* < 0.0001.

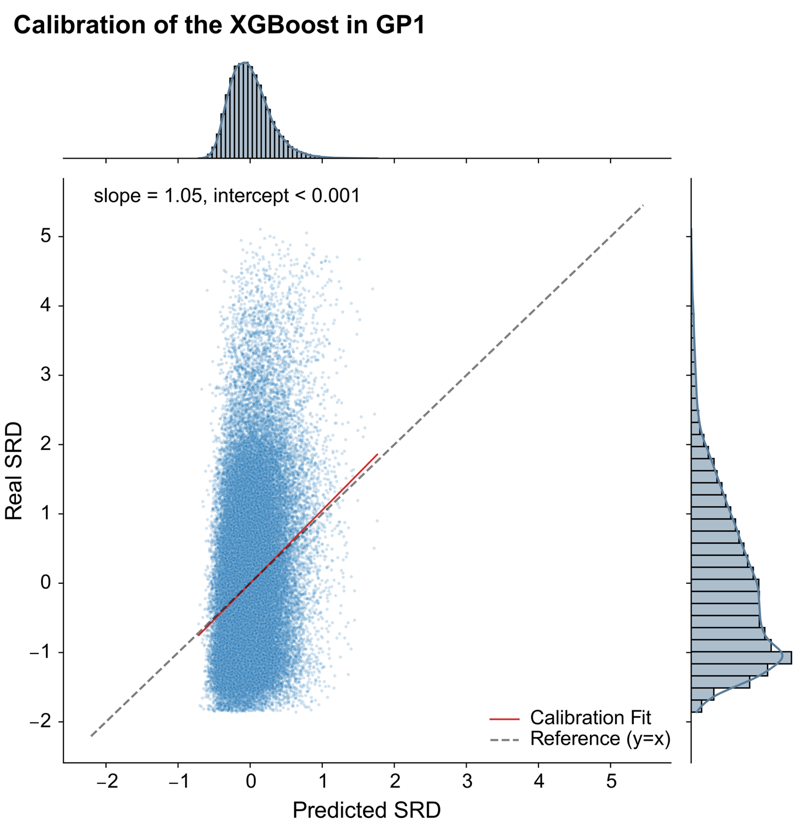

**Supplemental Figure 3: Calibration of XGBoost in GP1.** Each point represents an individual-level SRD score averaged across the test sets; model calibration is summarized by the calibration slope and intercept; histograms depict the distributions of predicted and observed SRD scores. The red solid line represents the fitted line, and the dashed line indicates perfect calibration (predicted SRD = real SRD). *GP: general population subsample; SRD: sleep-related depression; SHAP: SHapley Additive exPlanations.*

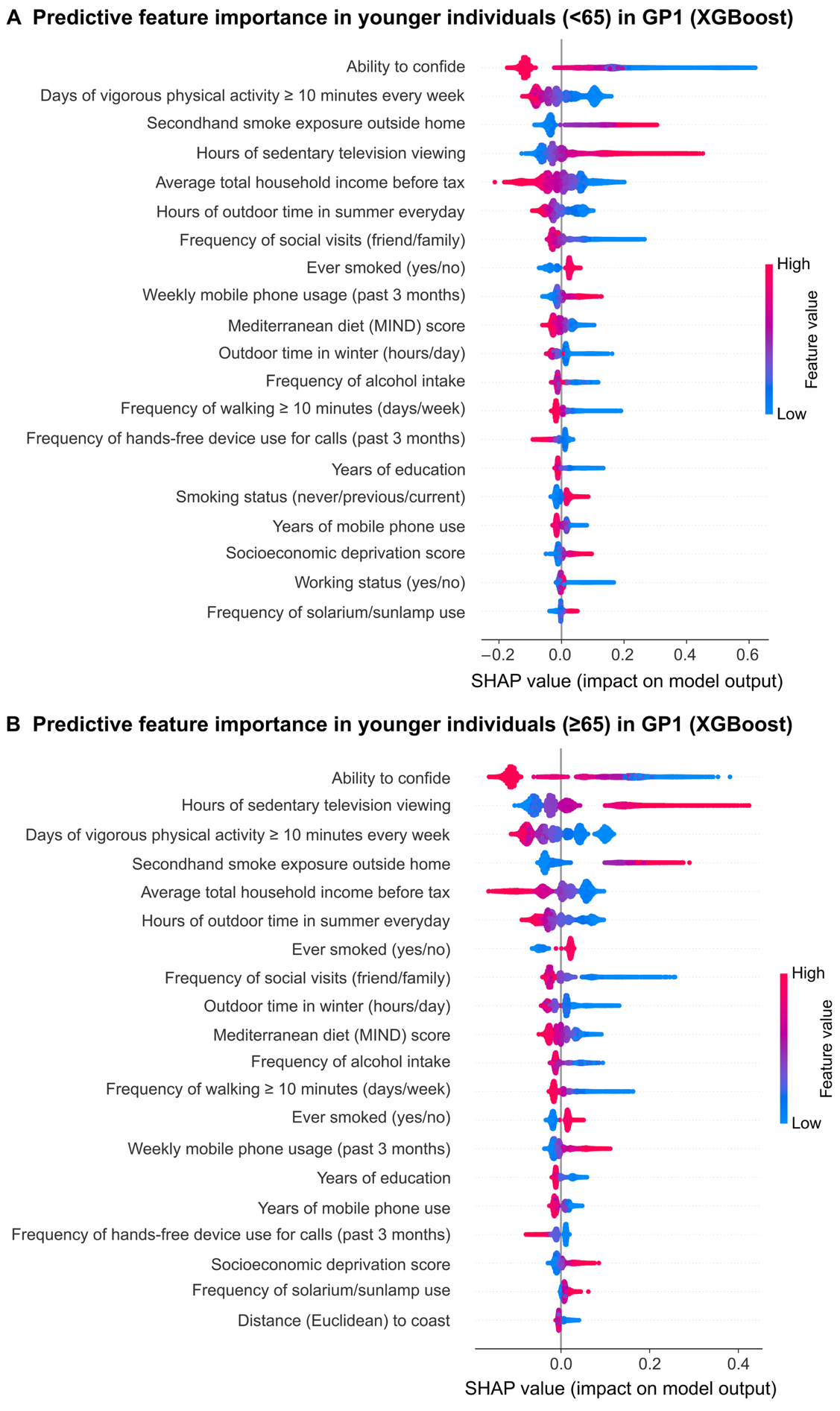

**Supplemental Figure 4. Feature importance for predicting sleep-related depression (SRD) across age-specific subgroups.** SHAP values indicate feature importance in the prediction models in younger (< 65 years; A) and older (≥ 65 years; B) participants in GP1. The x-axis denotes the direction and magnitude of the feature contribution to SRD prediction, where positive values reflect higher predicted SRD and negative values reflect lower predicted SRD. Color reflects feature values: blue indicates low and red indicates high feature scores. Only the top 20 features were visualized. *GP: general population; SHAP: SHapley Additive exPlanations*

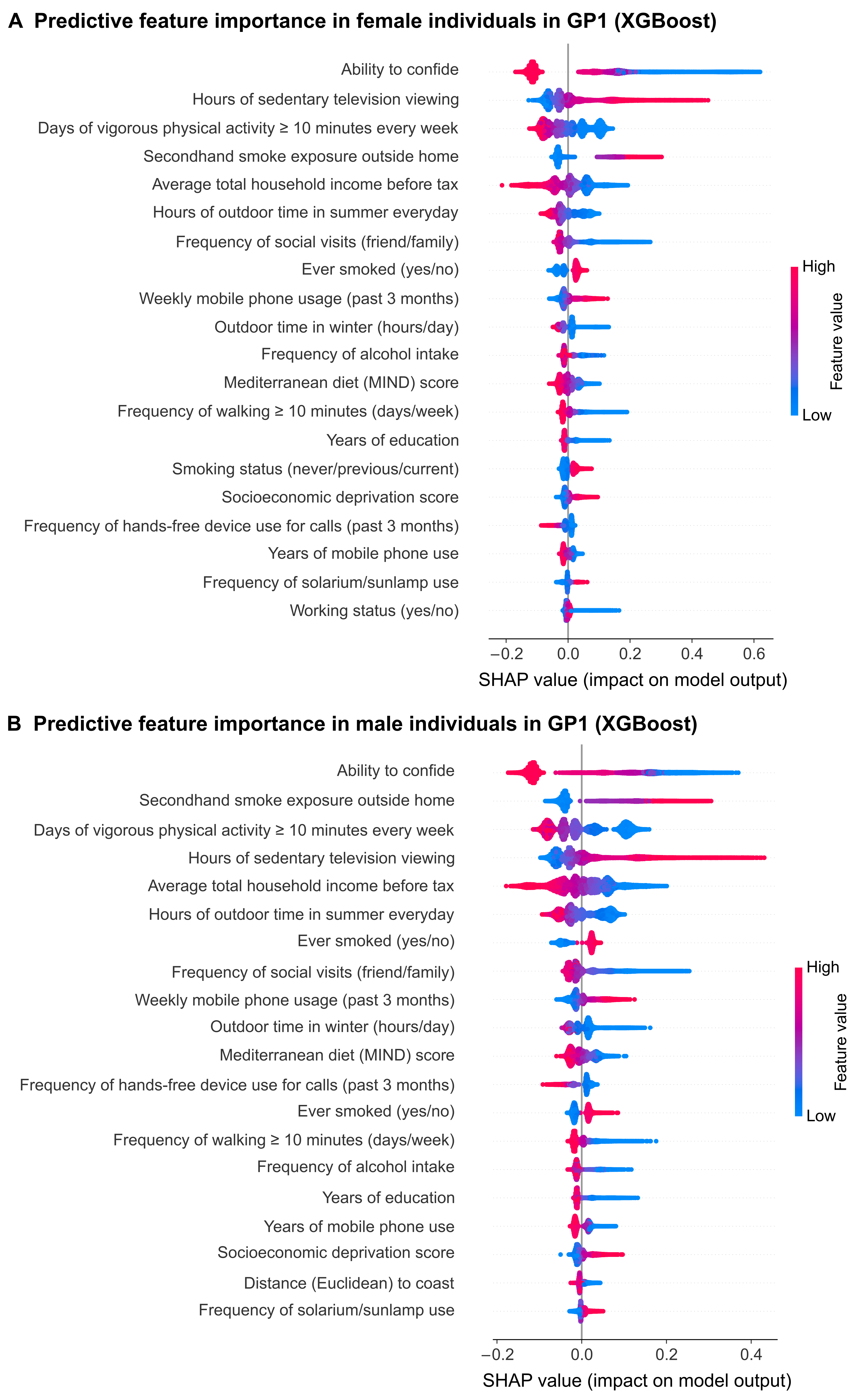

**Supplemental Figure 5. Feature importances for predicting sleep-related depression (SRD) across sex-specific subgroups.** SHAP values indicate feature importance in the prediction models in females (A) and males (B) in GP1. The x-axis denotes the direction and magnitude of the feature contribution to SRD prediction, where positive values reflect higher predicted SRD and negative values reflect lower predicted SRD. Color denotes feature values, with blue indicating low and red indicating high feature scores. Only the top 20 features were visualized. *GP: general population; SHAP: SHapley Additive exPlanations*

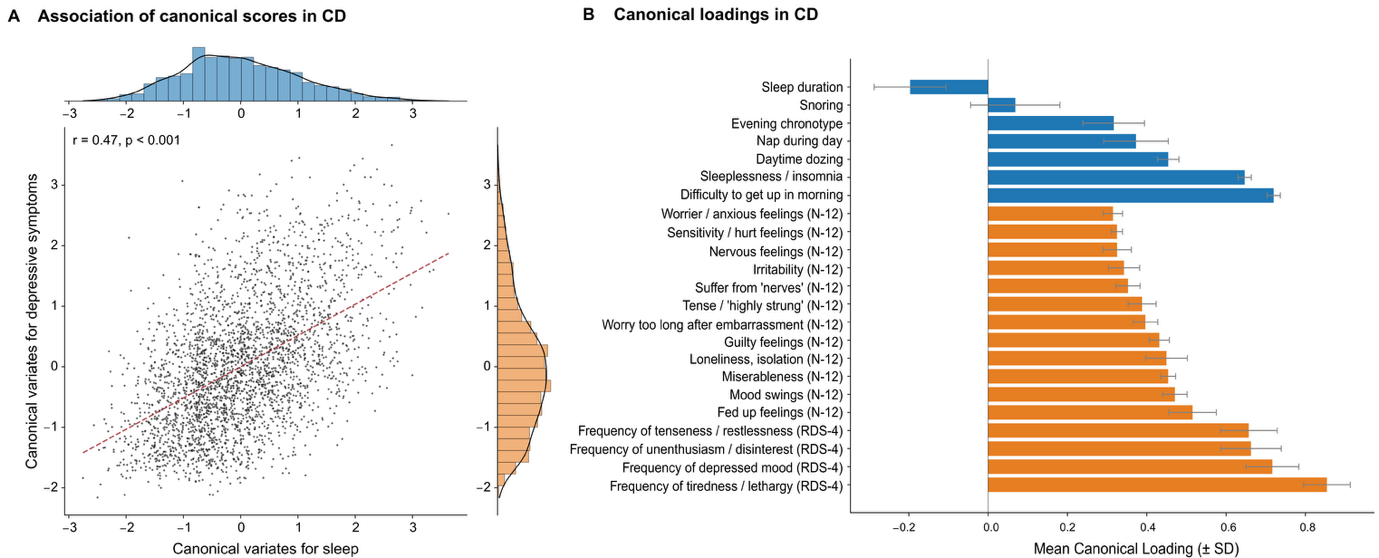

**Supplemental Figure 6: First significant latent dimension and loadings for sleep health and depression in clinical depression (CD). A.** Multivariate associations between canonical variates for sleep health (SH) and canonical variates of depression in CD. Each point represents an individual-level SRD score, where scores averaged across the test sets; model performance is illustrated using the correlation coefficient (r); blue and orange bar plots show the distributions of latent scores for SH and depression, respectively. **B.** Mean loadings across five outer CCA folds. Error bars indicate one standard deviation. Greater disturbances in SH loadings were associated with higher depression loadings. Blue represents SH, and orange reflects depression. *N-12: neuroticism-12; RDS-4: recent depressive symptoms, indicating depressive symptoms in the past two weeks; SRD: sleep-related depression.*

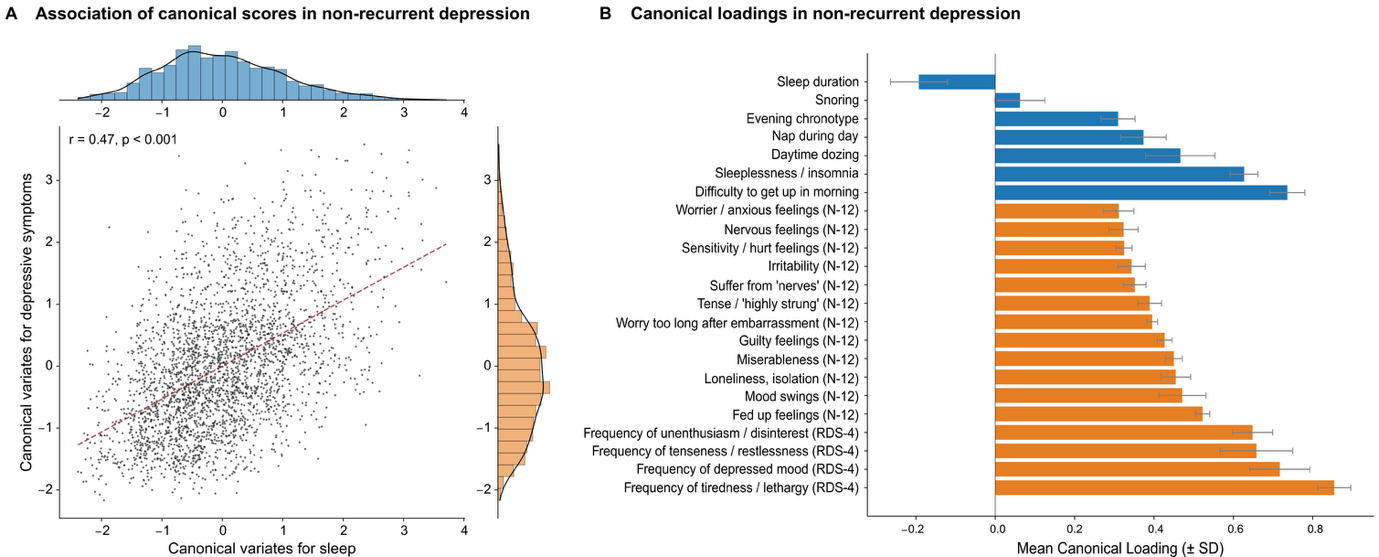

**Supplemental Figure 7:** **First significant latent dimension and loadings for sleep health and depression in clinical depression without recurrent depression. A.** Multivariate associations between canonical variates for sleep health (SH) and canonical variates of depression in non-recurrent depression. Each point represents an individual-level SRD score, where scores averaged across the test sets; model performance is illustrated using the correlation coefficient (r); blue and orange bar plots show the distributions of latent scores for SH and depression, respectively. **B.** Mean loadings across five outer CCA folds. Error bars indicate one standard deviation. Greater disturbances in SH loadings were associated with higher depression loadings. Blue represents SH, and orange reflects depression. *N-12: neuroticism-12; RDS-4: recent depressive symptoms, indicating depressive symptoms in the past two weeks; SRD: sleep-related depression.*

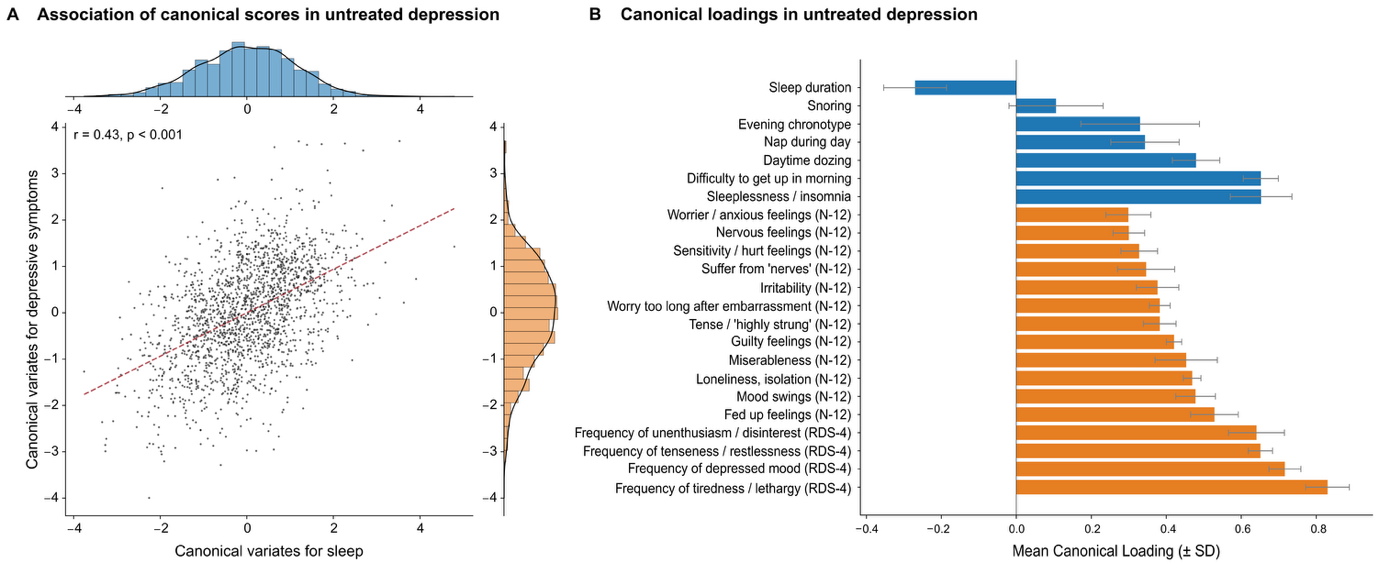

**Supplemental Figure 8: First significant latent dimension and loadings for sleep health and depression in untreated clinical depression. A.** Multivariate associations between canonical variates for sleep health (SH) and canonical variates of depression in individuals with untreated clinical depression. Each point represents an individual-level SRD score, where scores are averaged across the test sets; model performance is illustrated using the correlation coefficient (r); blue and orange bar plots show the distributions of latent scores for SH and depression, respectively. **B.** Mean loadings across five outer CCA folds. Error bars indicate one standard deviation. Greater disturbances in SH loadings were associated with higher depression loadings. Blue represents SH, and orange reflects depression. *N-12: neuroticism-12; RDS-4: recent depressive symptoms, indicating depressive symptoms in the past two weeks; SRD: sleep-related depression*

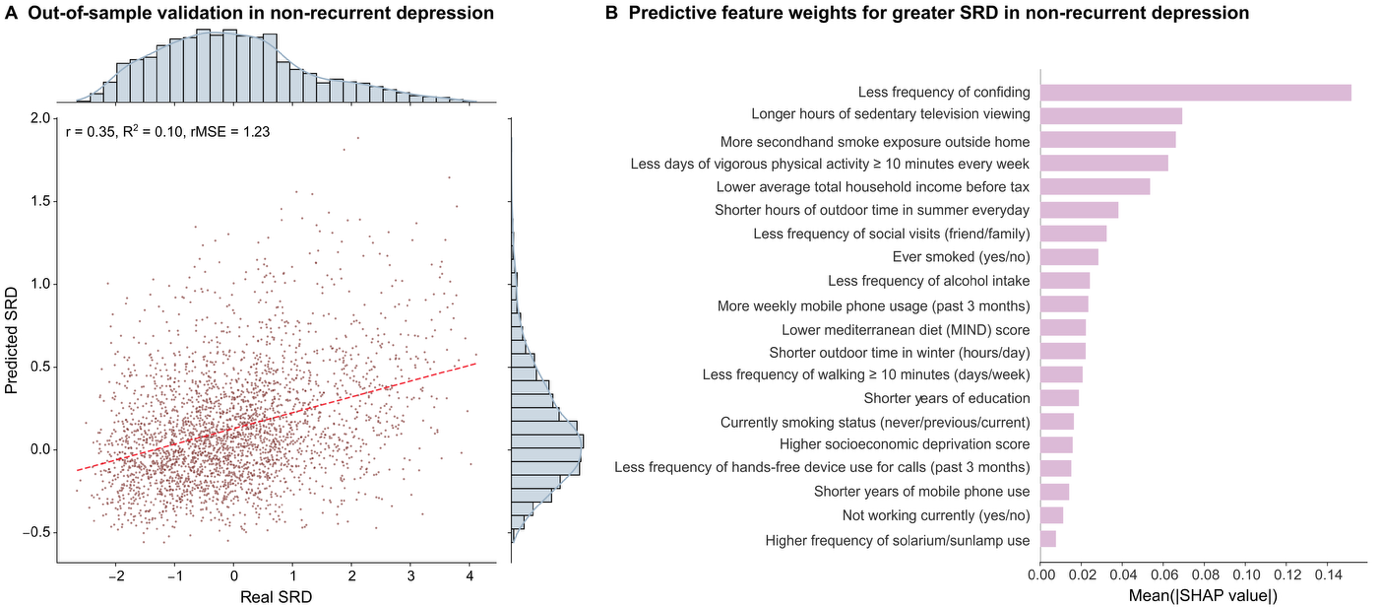

**Supplemental Figure 9: Predictive model performance in non-recurrent depression**. **A.** Each point represents an individual; bar plots represent the distribution of real and predicted SRD scores. Model performance is illustrated using the correlation coefficient (r), coefficient of determination (R²), and root mean square error (rMSE). **B.** Weights for each exposome factor in predicting higher SRD in GP1. The length of each bar represents the mean absolute SHAP values across the split test sets, reflecting the importance of each exposome factor in predicting higher SRD. Only the top 20 features were visualized. *SRD: sleep-related depressive symptoms; SHAP: SHapley Additive exPlanations.*

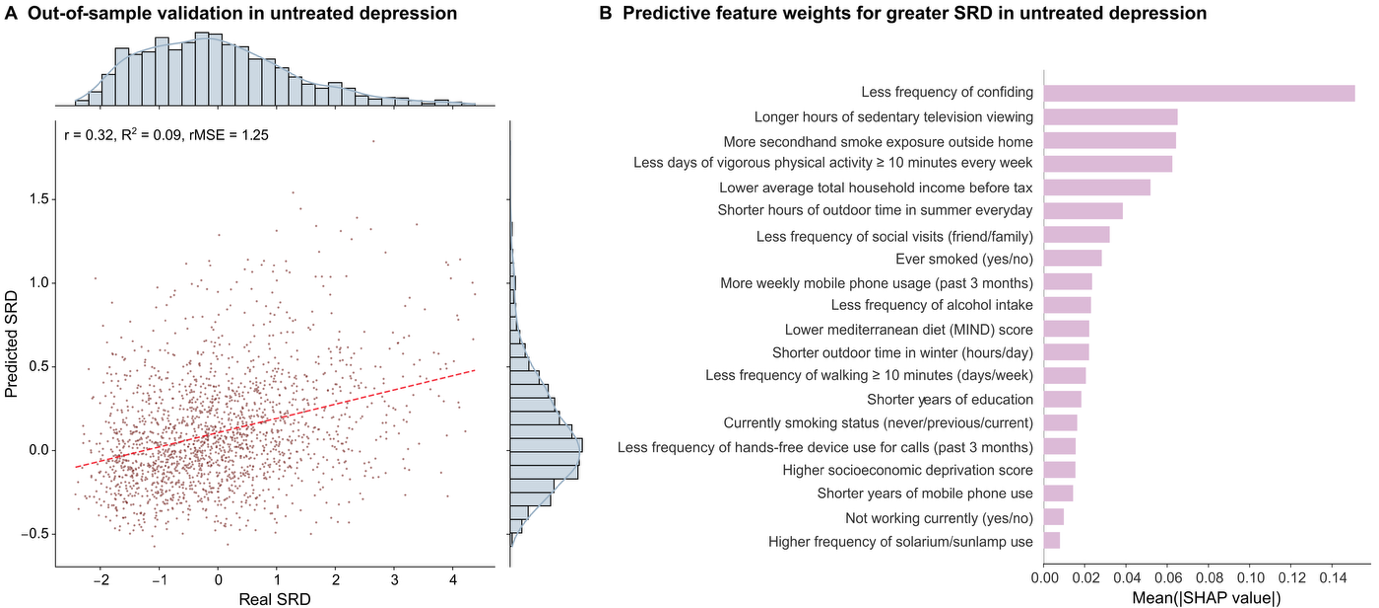

**Supplemental Figure 10: Predictive model performance in untreated depression**. **A.** Each point represents an individual; bar plots represent the distribution of real and predicted SRD scores. Model performance is illustrated using the correlation coefficient (r), coefficient of determination (R²), and root mean square error (rMSE). **B.** Weights for each exposome factor in predicting higher SRD in GP1. The length of each bar represents the mean absolute SHAP values across the split test sets, reflecting the importance of each exposome factor in predicting higher SRD. Only the top 20 features were visualized. *SRD: sleep-related depression; SHAP: SHapley Additive exPlanations.*

**Supplemental Table 1**

| **Detailed variables for exposome factors** | |
| --- | --- |
| **Catergory** | **Name in UKB** |
| Socioeconomics (economic deprivation) | 189 Townsend deprivation index at recruitment |
| Socioeconomics (household income) | 738 Average total household income before tax |
| Socioeconomics (social support) | 1031 Frequency of friend/family visits |
|  | 2110 Able to confide |
| Socioeconomics (working status) | 6142 Current employment status |
| Socioeconomics (education) | Years of education |
| Sleep health | 1160 Sleep duration |
|  | 1170 Getting up in morning |
|  | 1180 Morning/evening person (chronotype) |
|  | 1190 Nap during day |
|  | 1200 Sleeplessness / insomnia |
|  | 1210 Snoring |
|  | 1220 Daytime dozing / sleeping |
| Depressive symptoms (RDS-4) | 2050 Frequency of depressed mood in last 2 weeks |
|  | 2060 Frequency of unenthusiasm/disinterest in last 2 weeks |
|  | 2070 Frequency of tenseness/restlessness in last 2 weeks |
|  | 2080 Frequency of tiredness/lethargy in last 2 weeks |
| Depressive symptoms (N-12) | 1920 Mood swings |
|  | 1930 Miserableness |
|  | 1940 Irritability |
|  | 1950 Sensitivity/hurt feelings |
|  | 1960 Fed-up feelings |
|  | 1970 Nervous feelings |
|  | 1980 Worrier/anxious feelings |
|  | 1990 Tense/“highly strung” |
|  | 2000 Worry too long after embarrassment |
|  | 2010 Suffer from “nerves” |
|  | 2020 Loneliness, isolation |
|  | 2030 Guilty feelings |
| Lifestyle (diet) | 1289 Cooked vegetable intake |
|  | 1299 Salad / raw vegetable intake |
|  | 1309 Fresh fruit intake |
|  | 1319 Dried fruit intake |
|  | 1329 Oily fish intake |
|  | 1339 Non-oily fish intake |
|  | 1349 Processed meat intake |
|  | 1359 Poultry intake |
|  | 1369 Beef intake |
|  | 1379 Lamb/mutton intake |
|  | 1389 Pork intake |
|  | 1408 Cheese intake |
|  | 1428 Spread type |
|  | 1438 Bread intake |
|  | 1448 Bread type |
|  | 1458 Cereal intake |
|  | 1468 Cereal type |
|  | 1478 Salt added to food |
|  | 1488 Tea intake |
|  | 1498 Coffee intake |
|  | 1548 Variation in diet |
|  | 2654 Non-butter spread type details |
| Lifestyle (alcohol intake) | 20117 Alcohol drinker status |
|  | 1558 Alcohol intake frequency |
| Lifestyle (smoking) | 20160 Ever smoked |
|  | 20116 Smoking status |
|  | 1269 Exposure to tobacco smoke at home |
|  | 1279 Exposure to tobacco smoke outside home |
| Lifestyle (physical activity) | 884 Number of days/week of moderate physical activity 10+ minutes |
|  | 904 Number of days/week of vigorous physical activity 10+ minutes |
|  | 864 Number of days/week walked 10+ minutes |
|  | 1070 Time spent watching television (TV) |
| Lifestyle (digital use) | 1110 Length of mobile phone use |
|  | 1120 Weekly usage of mobile phone in last 3 months |
|  | 1130 Hands-free device/speakerphone use with mobile phone in last 3 month |
| Lifestyle (sun exposure) | 1050 Time spend outdoors in summer |
|  | 1060 Time spent outdoors in winter |
|  | 2277 Frequency of solarium/sunlamp use |
| Environment (air pollution) | 24014 Close to major road |
|  | 24012 Inverse distance to the nearest major road |
|  | 24010 Inverse distance to the nearest road |
|  | 24005 Particulate matter air pollution (pm10); 2010 |
|  | 24015 Sum of road length of major roads within 100m |
|  | 24011 Traffic intensity on the nearest major road |
|  | 24009 Traffic intensity on the nearest road |
|  | 24006 Particulate matter air pollution (pm2.5); 2010 |
| Environment (noise pollution) | 24024 Average 24-hour sound level of noise pollution |
| Environment (Greenspace and coastal proximity ) | 24508 Distance (Euclidean) to coast |
|  | 24503 Greenspace percentage, buffer 300m |
|  | 24502 Water percentage, buffer 1000m |
|  | 24505 Water percentage, buffer 300m |

**Supplemental Table 2:**

| **Exclusion crietria** | |
| --- | --- |
| **Catergory** | **Name in UKB** |
| ICD-10 | A80 Acute poliomyelitis |
|  | A81 Atypical virus infections of central nervous system |
|  | A83 Mosquito-borne viral encephalitis |
|  | A84 Tick-borne viral encephalitis |
|  | A85 Other viral encephalitis, not elsewhere classified |
|  | A86 Unspecified viral encephalitis |
|  | A87 Viral meningitis |
|  | A88 Other viral infections of central nervous system, not elsewhere classified |
|  | A89 Unspecified viral infection of central nervous system |
|  | F00 Dementia in Alzheimer's disease |
|  | F01 Vascular dementia |
|  | F02 Dementia in other diseases classified elsewhere |
|  | F03 Unspecified dementia |
|  | F04 Organic amnesic syndrome, not induced by alcohol and other psychoactive substances |
|  | F05 Delirium, not induced by alcohol and other psychoactive substances |
|  | F06 Other mental disorders due to brain damage and dysfunction and to physical disease |
|  | F07 Personality and behavioural disorders due to brain disease, damage and dysfunction |
|  | F09 Unspecified organic or symptomatic mental disorder |
|  | F10 Mental and behavioural disorders due to use of alcohol |
|  | F11 Mental and behavioural disorders due to use of opioids |
|  | F12 Mental and behavioural disorders due to use of cannabinoids |
|  | F13 Mental and behavioural disorders due to use of sedatives or hypnotics |
|  | F14 Mental and behavioural disorders due to use of cocaine |
|  | F15 Mental and behavioural disorders due to use of other stimulants, including caffeine |
|  | F16 Mental and behavioural disorders due to use of hallucinogens |
|  | F17 Mental and behavioural disorders due to use of tobacco |
|  | F18 Mental and behavioural disorders due to use of volatile solvents |
|  | F19 Mental and behavioural disorders due to multiple drug use and use of other psychoactive substances |
|  | F20 Schizophrenia |
|  | F21 Schizotypal disorder |
|  | F22 Persistent delusional disorders |
|  | F23 Acute and transient psychotic disorders |
|  | F24 Induced delusional disorder |
|  | F25 Schizoaffective disorders |
|  | F28 Other nonorganic psychotic disorders |
|  | F29 Unspecified nonorganic psychosis |
|  | F30 Manic episode |
|  | F31 Bipolar affective disorder |
|  | G00 Bacterial meningitis, not elsewhere classified |
|  | G02 Meningitis in other infectious and parasitic diseases classified elsewhere |
|  | G03 Meningitis due to other and unspecified causes |
|  | G04 Encephalitis, myelitis and encephalomyelitis |
|  | G05 Encephalitis, myelitis and encephalomyelitis in diseases classified elsewhere |
|  | G30 Alzheimer's disease |
|  | G31 Other degenerative diseases of nervous system, not elsewhere classified |
|  | G32 ther degenerative disorders of nervous system in diseases classified elsewhere |
|  | G40 Epilepsy |
|  | G41 Status epilepticus |
|  | G46 Vascular syndromes of brain in cerebrovascular diseases |
|  | G80 Infantile cerebral palsy |
|  | G81 Hemiplegia |
|  | G82 Paraplegia and tetraplegia |
|  | G83 Other paralytic syndromes |
|  | S01 Open wound of head |
|  | S04 Injury of cranial nerves |
|  | S06 Intracranial injury |
|  | S07 Crushing injury of head |
|  | S08 Traumatic amputation of part of head |
|  | S09 Other and unspecified injuries of head |
|  | F70 Mild mental retardation |
|  | F71 Moderate mental retardation |
|  | F72 Severe mental retardation |
|  | F78 Other mental retardation |
|  | F79 Unspecified mental retardation |
| Self-reported from Data-Coding 6 | 1031 meningeal cancer / malignant meningioma |
|  | 1032 brain cancer / primary malignant brain tumour |
|  | 1081 stroke |
|  | 1082 transient ischaemic attack (TIA) |
|  | 1083 subdural haemorrhage/haematoma |
|  | 1086 subarachnoid haemorrhage |
|  | 1123 sleep apnoea |
|  | 1240 neurological injury/trauma |
|  | 1244 infection of nervous system |
|  | 1245 brain abscess / intracranial abscess |
|  | 1246 encephalitis |
|  | 1247 meningitis |
|  | 1258 chronic/degenerative neurological problem |
|  | 1259 motor neurone disease |
|  | 1261 multiple sclerosis |
|  | 1262 parkinsons disease |
|  | 1263 dementia/alzheimers/cognitive impairment |
|  | 1264 epilepsy |
|  | 1266 head injury |
|  | 1289 schizophrenia |
|  | 1291 mania/bipolar disorder/manic depression |
|  | 1397 other demyelinating disease (not multiple sclerosis) |
|  | 1409 opioid dependency |
|  | 1410 other substance abuse/dependency |
|  | 1433 cerebral palsy |
|  | 1434 other neurological problem |
|  | 1491 brain haemorrhage |
|  | 1524 spina bifida |
|  | 1583 ischaemic stroke |
|  | 1626 fracture skull / head |
|  | 1659 meningioma / benign meningeal tumour |
|  | 1683 benign neuroma |

**Supplemental Table 3**

| **Medication codes from Data-Coding 4 in the UK Biobank** | |
| --- | --- |
| Codes for antipsychotics | 1140868170, 1140928916, 1141152848, 1140867444, 1140879658, 1140868120, 1141153490, 1140867304, 1141152860, 1140867168, 1141195974, 1140867244, 1140867152, 1140909800, 1140867420, 1140879746, 1141177762, 1140867456, 1140867952, 1140867150, 1141167976, 1140882100, 1140867342, 1140863416, 1141202024, 1140882098, 1140867184, 1140867092, 1140882320, 1140910358, 1140867208, 1140909802, 1140867134, 1140867306, 1140867210, 1140867398, 1140867078, 1140867218, 1141201792, 1141200458, 1140867136, 1140879750, 1140867180, 1140867546, 1140928260, 1140927956. |
| Codes for antidepressants | 1140879616, 1140921600, 1140879540, 1140867878, 1140916282, 1140909806, 1140867888, 1141152732, 1141180212, 1140879634, 1140867876, 1140882236, 1141190158, 1141200564, 1140867726, 1140879620, 1140867818, 1140879630, 1140879628, 1141151946, 1140867948, 1140867624, 1140867756, 1140867884, 1141151978, 1141152736, 1141201834, 1140867690, 1140867640, 1140867920, 1140867850, 1140879544, 1141200570, 1140867934, 1140867758, 1140867914, 1140867820, 1141151982, 1140882244, 1140879556, 1140867852, 1140867860, 1140917460, 1140867938, 1140867856, 1140867922, 1140910820, 1140882312, 1140867944, 1140867784, 1140867812, 1140867668. |
| Data-Coding 4 lists the codes recording medical treatments used by clinical nurses (https://biobank.ndph.ox.ac.uk/ukb/coding.cgi?id=4&nl=1). | |

**Supplemental Table 4**

| **Sample attrition by data category** | | | | |
| --- | --- | --- | --- | --- |
| Exclusion criterion | GP1: N excluded | GP1: N remaining | CD: N excluded | CD: N remaining |
| Initial sample after exclusion | - | 386,543 | - | 17,043 |
| Missing demographic variables | 0 | 386,543 | 0 | 17,043 |
| Missing socioeconomic variables | 58,556 | 327,987 | 2,974 | 14,069 |
| Missing depressive symptom variables | 70,608 | 257,379 | 3,709 | 10,360 |
| Missing sleep health variables | 33,631 | 223,748 | 1,474 | 8,886 |
| Missing working status variables | 809 | 222,939 | 42 | 8,844 |
| Missing environmental variables | 31,946 | 190,993 | 551 | 8,293 |
| Missing social support variables | 2,978 | 188,015 | 97 | 8,196 |
| Missing lifestyle variables | 115,095 | 72,920 | 5,150 | 3,046 |
| Abbreviation: GP1: general population 1; CD: clinical depression | | | | |

**Supplemental Table 5**

| **Clinical subsamples for sensitivity analysis** | | | | |
| --- | --- | --- | --- | --- |
| **Subsamples** | **Sample** | **Age, mean [SE]** | **Sex, (females%)** | **Definition** |
| Non-recurrent depression | 2,969 | 55.77 [7.93] | 66.9% | Recurrent depression was excluded from the clinical depression group, regardless of antidepressant use. |
| Un-treated depression | 1,863 | 55.68 [8.10] | 64.6% | Both individuals with recurrent depression and those using antidepressants while recruited were excluded. |

**Supplemental Table 6**

| **Demographic characteristics and feature distributions across data domains** | | | | |
| --- | --- | --- | --- | --- |
|  | **GP1 (n = 64,781)** | **GP2 at baseline  (n = 8,139)** | **GP2 at follow-up (n = 8,139)** | **CD (n = 3,046)** |
| **Demographic characteristics** |  |  |  |  |
| Age, mean [SE], y | 55.83 [8.10] | 54.76 [7.50] | 64.01 [7.66] | 55.79 [7.93] |
| Sex, No. (%) |  |  |  |  |
| Females | 50.05% | 45.44% | 45.44% | 66.84% |
| **Features** |  |  |  |  |
| Years of education | 15.06 [4.57] | 16.76 [3.68] | - | 14.52 [4.81] |
| Average total household income before tax | 3.0 [2.0, 4.0] | 3.0 [2.0, 4.0] | - | 2.0 [1.0, 3.0] |
| Socioeconomic deprivation score | -1.80 [2.76] | -2.13 [2.57] | - | -1.30 [3.03] |
| Frequency of alcohol intake | 4.0 [4.0, 5.0] | 5.0 [4.0, 5.0] | - | 4.0 [3.0, 5.0] |
| Alcohol drinking status (never/previous/current) | 2.20% : 2.81% : 94.98% | 1.71% : 1.50% : 96.79% | - | 4.10% : 5.29% : 90.61% |
| Ever smoked (yes/no) | 57.08% : 42.92% | 55.31% : 44.69% | - | 59.03% : 40.97% |
| Secondhand smoke exposure outside home | 0.45 [2.12] | 0.34 [1.57] | - | 0.78 [3.87] |
| Smoking status (never/previous/current) | 59.00% : 38.49% : 2.51% | 62.07% : 35.91% : 2.01% | - | 54.99% : 42.88% : 2.13% |
| Secondhand smoke exposure at home | 0.40 [3.66] | 0.32 [3.47] | - | 0.76 [5.26] |
| MIND diet score | 6.91 [1.06] | 6.94 [1.05] | - | 6.87 [1.15] |
| Frequency of walking ≥ 10 minutes (days/week) | 5.57 [1.82] | 5.46 [1.88] | - | 5.46 [1.98] |
| Days of moderate physical activity ≥ 10 minutes every week | 3.79 [2.26] | 3.65 [2.21] | - | 3.66 [2.35] |
| Days of vigorous physical activity ≥ 10 minutes/week | 2.07 [1.94] | 2.05 [1.84] | - | 1.78 [1.95] |
| Hours of sedentary television viewing | 2.69 [1.41] | 2.48 [1.31] | - | 2.99 [1.59] |
| Frequency of confiding | 5.0 [3.0, 5.0] | 5.0 [3.0, 5.0] | - | 5.0 [2.0, 5.0] |
| Frequency of social visits (friend/family) | 5.0 [5.0, 6.0] | 5.0 [5.0, 6.0] | - | 5.0 [5.0, 6.0] |
| Weekly mobile phone usage (past 3 months) | 1.0 [1.0, 2.0] | 1.0 [1.0, 2.0] | - | 1.0 [1.0, 2.0] |
| Frequency of hands-free device use for calls (past 3 months) | 0.0 [0.0, 0.0] | 0.0 [0.0, 0.0] | - | 0.0 [0.0, 0.0] |
| Years of mobile phone use | 3.0 [3.0, 4.0] | 3.0 [3.0, 4.0] | - | 3.0 [3.0, 4.0] |
| Hours of outdoor time in winter everyday | 2.16 [1.77] | 1.93 [1.53] | - | 2.11 [1.75] |
| Hours of outdoor time in summer every day | 4.09 [2.30] | 3.79 [2.15] | - | 4.04 [2.22] |
| Frequency of solarium/sunlamp use | 0.60 [5.93] | 0.63 [3.94] | - | 1.00 [6.16] |
| Working status (yes/no) | 67.65% : 32.35% | 74.09% : 25.91% | - | 60.24% : 39.76% |
| Close to major road (yes/no) | 6.78% : 93.22% | 7.36% : 92.64% | - | 6.63% : 93.37% |
| Inverse distance to the nearest major road | 0.01 [0.02] | 0.01 [0.02] | - | 0.01 [0.01] |
| Inverse distance to the nearest road | 0.05 [0.07] | 0.05 [0.07] | - | 0.05 [0.09] |
| Particulate matter air pollution (pm10) in 2010 | 16.13 [1.89] | 15.98 [1.89] | - | 16.11 [1.83] |
| Sum of road length of major roads within 100m | 26.37 [73.54] | 28.91 [77.21] | - | 25.95 [74.49] |
| Traffic intensity on the nearest major road | 23177.90 [21158.61] | 22229.24 [20429.16] | - | 22599.63 [19781.02] |
| Traffic intensity on the nearest road | 1497.07 [4902.63] | 1543.36 [5017.70] | - | 1364.18 [4217.96] |
| Particulate matter air pollution (pm2.5) in 2010 | 9.88 [1.01] | 9.87 [1.04] | - | 9.97 [1.01] |
| Average 24-hour sound level of noise pollution | 55.96 [4.21] | 56.03 [4.29] | - | 56.00 [4.18] |
| Distance (Euclidean) to coast | 45.30 [27.08] | 45.88 [27.08] | - | 44.03 [27.78] |
| Greenspace_percentage, buffer 300m | 36.77 [23.97] | 38.99 [24.58] | - | 36.18 [22.42] |
| Water_percentage, buffer 1000m | 1.26 [2.46] | 1.28 [2.59] | - | 1.22 [2.51] |
| Water_percentage, buffer 300m | 0.91 [3.00] | 0.96 [3.18] | - | 0.87 [3.00] |
| Continuous variables are presented as mean [SD], ordinal variables as median [IQR], and categorical variables as percentage. GP: General population; CD: clinically diagnosed depression; SD: standard deviation; IQR: interquartile range. Features for GP2 at folow-up were from GP2 at baselne. | | | | |

**Supplemental Table 7**

| **Variance in depressive symptoms and SH explained by opposing canonical variates across folds** | | | | | | |
| --- | --- | --- | --- | --- | --- | --- |
| **Samples** | **Variables** | **Fold 1** | **Fold 2** | **Fold 3** | **Fold 4** | **Fold 5** |
| GP1 | SH | 18.45% | 18.14% | 18.61% | 18.35% | 18.33% |
|  | Depression | 23.36% | 23.87% | 24.96% | 24.21% | 24.65% |
| CD | SH | 21.61% | 21.52% | 20.82% | 21.41% | 18.94% |
|  | Depression | 28.34% | 26.68% | 26.02% | 22.93% | 23.86% |
| *Abbreviation: SH, sleep health; CD: clinical depression subsamples; GP: general population subsamples* | | | | | | |

**Supplemental Table 8**

| **Variance inflation factor (VIF) scores for input features in predictive model** | | | | | |
| --- | --- | --- | --- | --- | --- |
| **Features** | **VIF fold 1** | **VIF fold 2** | **VIF fold 3** | **VIF fold 4** | **VIF fold 5** |
| Particulate matter air pollution (pm10); 2010 | 3.78 | 3.76 | 3.80 | 3.82 | 3.81 |
| Close to major road | 3.47 | 3.47 | 3.53 | 3.43 | 3.44 |
| Average 24-hour sound level of noise pollution | 3.33 | 3.39 | 3.44 | 3.36 | 3.35 |
| Particulate matter air pollution (pm2.5); 2010 | 2.83 | 2.85 | 2.86 | 2.85 | 2.86 |
| Sum of road length of major roads within 100m | 2.82 | 2.81 | 2.85 | 2.84 | 2.80 |
| Traffic intensity on the nearest major road | 2.55 | 2.53 | 2.55 | 2.57 | 2.56 |
| Greenspace percentage, buffer 300m | 2.31 | 2.31 | 2.32 | 2.31 | 2.33 |
| Water percentage, buffer 300m | 2.13 | 2.06 | 2.11 | 2.13 | 2.13 |
| Water percentage, buffer 1000m | 2.11 | 2.04 | 2.10 | 2.11 | 2.11 |
| Time spend outdoors in summer | 1.97 | 1.96 | 1.95 | 1.96 | 1.97 |
| Time spent outdoors in winter | 1.96 | 1.96 | 1.95 | 1.96 | 1.96 |
| Smoking status | 1.96 | 1.96 | 1.96 | 1.96 | 1.95 |
| Ever smoked | 1.95 | 1.96 | 1.95 | 1.95 | 1.94 |
| Average total household income before tax | 1.53 | 1.53 | 1.52 | 1.52 | 1.53 |
| Inverse distance to the nearest major road | 1.52 | 1.56 | 1.61 | 1.56 | 1.57 |
| Number of days/week of moderate physical activity 10+ minutes | 1.49 | 1.48 | 1.49 | 1.49 | 1.49 |
| Alcohol intake frequency | 1.47 | 1.47 | 1.46 | 1.46 | 1.46 |
| Traffic intensity on the nearest road | 1.45 | 1.44 | 1.45 | 1.42 | 1.43 |
| Inverse distance to the nearest road | 1.37 | 1.37 | 1.36 | 1.36 | 1.36 |
| Alcohol drinker status | 1.37 | 1.36 | 1.36 | 1.36 | 1.36 |
| Number of days/week of vigorous physical activity 10+ minutes | 1.36 | 1.36 | 1.37 | 1.37 | 1.37 |
| Current employment status | 1.36 | 1.36 | 1.35 | 1.35 | 1.36 |
| Weekly usage of mobile phone in last 3 months | 1.30 | 1.29 | 1.30 | 1.29 | 1.30 |
| Townsend deprivation index at recruitment | 1.29 | 1.30 | 1.30 | 1.30 | 1.30 |
| Years of education | 1.19 | 1.19 | 1.19 | 1.19 | 1.19 |
| Hands-free device/speakerphone use with mobile phone in last 3 month | 1.19 | 1.19 | 1.19 | 1.19 | 1.19 |
| Number of days/week walked 10+ minutes | 1.18 | 1.17 | 1.18 | 1.18 | 1.18 |
| Time spent watching television (TV) | 1.17 | 1.18 | 1.17 | 1.18 | 1.17 |
| Length of mobile phone use | 1.17 | 1.17 | 1.17 | 1.17 | 1.17 |
| Frequency of friend/family visits | 1.08 | 1.07 | 1.07 | 1.07 | 1.07 |
| Distance (Euclidean) to coast | 1.07 | 1.07 | 1.07 | 1.07 | 1.07 |
| Exposure to tobacco smoke outside home | 1.05 | 1.05 | 1.06 | 1.05 | 1.05 |
| Exposure to tobacco smoke at home | 1.03 | 1.03 | 1.04 | 1.03 | 1.03 |
| Able to confide | 1.03 | 1.03 | 1.03 | 1.03 | 1.03 |
| Mediterranean diet (MIND) scores | 1.02 | 1.02 | 1.02 | 1.02 | 1.02 |
| Frequency of solarium/sunlamp use | 1.01 | 1.01 | 1.01 | 1.01 | 1.01 |

**Supplemental Table 8**

| **Group comparison of sex for SHAP values from XGBoost in GP 1** | | | |
| --- | --- | --- | --- |
| **Features** | **t stat** | **p value** | **p fdr** |
| Years of education | 2.93 | 0.003 | 0.005 |
| Townsend deprivation index at recruitment | -0.45 | 0.653 | 0.653 |
| Close to major road | -96.72 | < 0.001 | < 0.001 |
| Inverse distance to the nearest major road | -6.11 | < 0.001 | < 0.001 |
| Inverse distance to the nearest road | -16.68 | < 0.001 | < 0.001 |
| Particulate matter air pollution (pm10); 2010 | 2.86 | 0.004 | 0.006 |
| Sum of road length of major roads within 100m | 2.47 | 0.014 | 0.018 |
| Traffic intensity on the nearest major road | 2.29 | 0.022 | 0.027 |
| Traffic intensity on the nearest road | 17.90 | < 0.001 | < 0.001 |
| Particulate matter air pollution (pm2.5); 2010 | 2.29 | 0.022 | 0.027 |
| Average 24-hour sound level of noise pollution | 1.39 | 0.164 | 0.169 |
| Distance (Euclidean) to coast | 4.11 | < 0.001 | < 0.001 |
| Greenspace percentage, buffer 300m | -3.79 | < 0.001 | < 0.001 |
| Water percentage, buffer 1000m | 6.15 | < 0.001 | < 0.001 |
| Water percentage, buffer 300m | 54.30 | < 0.001 | < 0.001 |
| Time spend outdoors in summer | -12.34 | < 0.001 | < 0.001 |
| Time spent outdoors in winter | 1.96 | 0.050 | 0.055 |
| Frequency of solarium/sunlamp use | 100.23 | < 0.001 | < 0.001 |
| Average total household income before tax | 10.79 | < 0.001 | < 0.001 |
| Alcohol intake frequency | 9.52 | < 0.001 | < 0.001 |
| Alcohol drinker status | -1.61 | 0.108 | 0.114 |
| Ever smoked | -10.53 | < 0.001 | < 0.001 |
| Smoking status | -9.74 | < 0.001 | < 0.001 |
| Exposure to tobacco smoke at home | 28.60 | < 0.001 | < 0.001 |
| Exposure to tobacco smoke outside home | 8.44 | < 0.001 | < 0.001 |
| Current employment status | -32.54 | < 0.001 | < 0.001 |
| Hands-free device/speakerphone use with mobile phone in last 3 month | -15.55 | < 0.001 | < 0.001 |
| Weekly usage of mobile phone in last 3 months | -6.14 | < 0.001 | < 0.001 |
| Length of mobile phone use | -2.11 | 0.035 | 0.040 |
| Number of days/week walked 10+ minutes | -2.70 | 0.007 | 0.009 |
| Number of days/week of moderate physical activity 10+ minutes | 8.24 | < 0.001 | < 0.001 |
| Number of days/week of vigorous physical activity 10+ minutes | -6.66 | < 0.001 | < 0.001 |
| Time spent watching television (TV) | 1.97 | 0.049 | 0.055 |
| Frequency of friend/family visits | -3.58 | < 0.001 | < 0.001 |
| Able to confide | -4.85 | < 0.001 | < 0.001 |
| Mediterranean diet (MIND) scores | 6.20 | < 0.001 | < 0.001 |
| *Abbreviation: GP, general population; SHAP: SHapley Additive exPlanations; XGBoost: extreme gradient boosting* | | | |

**Supplemental Table 9**

| **Group comparison of age for SHAP values from XGBoost in GP 1** | | | |
| --- | --- | --- | --- |
| **Variable** | **t stat** | **p value** | **p fdr** |
| Years of education | 2.93 | 0.003 | 0.005 |
| Townsend deprivation index at recruitment | -0.45 | 0.653 | 0.653 |
| Close to major road | -96.72 | < 0.001 | < 0.001 |
| Inverse distance to the nearest major road | -6.11 | < 0.001 | < 0.001 |
| Inverse distance to the nearest road | -16.68 | < 0.001 | < 0.001 |
| Particulate matter air pollution (pm10); 2010 | 2.86 | 0.004 | 0.006 |
| Sum of road length of major roads within 100m | 2.47 | 0.014 | 0.018 |
| Traffic intensity on the nearest major road | 2.29 | 0.022 | 0.027 |
| Traffic intensity on the nearest road | 17.90 | < 0.001 | < 0.001 |
| Particulate matter air pollution (pm2.5); 2010 | 2.29 | 0.022 | 0.027 |
| Average 24-hour sound level of noise pollution | 1.39 | 0.164 | 0.169 |
| Distance (Euclidean) to coast | 4.11 | < 0.001 | < 0.001 |
| Greenspace percentage, buffer 300m | -3.79 | < 0.001 | < 0.001 |
| Water percentage, buffer 1000m | 6.15 | < 0.001 | < 0.001 |
| Water percentage, buffer 300m | 54.30 | < 0.001 | < 0.001 |
| Time spend outdoors in summer | -12.34 | < 0.001 | < 0.001 |
| Time spent outdoors in winter | 1.96 | 0.050 | 0.055 |
| Frequency of solarium/sunlamp use | 100.23 | < 0.001 | < 0.001 |
| Average total household income before tax | 10.79 | < 0.001 | < 0.001 |
| Alcohol intake frequency | 9.52 | < 0.001 | < 0.001 |
| Alcohol drinker status | -1.61 | 0.108 | 0.114 |
| Ever smoked | -10.53 | < 0.001 | < 0.001 |
| Smoking status | -9.74 | < 0.001 | < 0.001 |
| Exposure to tobacco smoke at home | 28.60 | < 0.001 | < 0.001 |
| Exposure to tobacco smoke outside home | 8.44 | < 0.001 | < 0.001 |
| Current employment status | -32.54 | < 0.001 | < 0.001 |
| Hands-free device/speakerphone use with mobile phone in last 3 month | -15.55 | < 0.001 | < 0.001 |
| Weekly usage of mobile phone in last 3 months | -6.14 | < 0.001 | < 0.001 |
| Length of mobile phone use | -2.11 | 0.035 | 0.040 |
| Number of days/week walked 10+ minutes | -2.70 | 0.007 | 0.009 |
| Number of days/week of moderate physical activity 10+ minutes | 8.24 | < 0.001 | < 0.001 |
| Number of days/week of vigorous physical activity 10+ minutes | -6.66 | < 0.001 | < 0.001 |
| Time spent watching television (TV) | 1.97 | 0.049 | 0.055 |
| Frequency of friend/family visits | -3.58 | < 0.001 | 0.001 |
| Able to confide | -4.85 | < 0.001 | < 0.001 |
| Mediterranean diet (MIND) scores | 6.20 | < 0.001 | < 0.001 |
| *Abbreviation:* *GP, general population; SHAP: SHapley Additive exPlanations; XGBoost: extreme gradient boosting* | | | |

**Supplemental Table 10**

| **Comparison of the full model and demographic model in predicting SRD** | | | |
| --- | --- | --- | --- |
| **Model** | **r (mean±SD)** | **R^2^ (mean±SD)** | **rMSE (mean±SD)** |
| Full model | 0.25 ± 0.011 | 0.063± 0.006 | 1.11 ± 0.014 |
| Demographic model | 0.02 ± 0.009 | 0.00 ± 0.001 | 1.14 ± 0.016 |
| ∆ (Full - Demographic) | 0.23 ± 0.015 | 0.06 ± 0.006 | -0.04 ± 0.004 |
| Full model used the same exposome factors as features; Demographic model used age, sex, and years of education as features. *SRD: sleep-related depression*. | | | |

**Supplemental Table 11**

| **Comparison of predictive performance with and without confounds in predicting SRD** | | | |
| --- | --- | --- | --- |
| **Model** | **r (mean±SD)** | **R^2^ (mean±SD)** | **rMSE (mean±SD)** |
| With confounds | 0.25 ± 0.011 | 0.06 ± 0.006 | 1.11 ± 0.014 |
| Without confounds | 0.25 ± 0.012 | 0.06 ± 0.006 | 1.11 ± 0.015 |
| Both model used the same exposome factors as features. With confounds model controlled age and sex as confounds, whereas without confounds model didn't control the prior confounds. *SRD: sleep-related depression.* | | | |

**Supplemental Table 12**

| **Model performance in out-of-sample validation** | | | |
| --- | --- | --- | --- |
| **Subsamples** | **r (mean±SD) [95% CI]** | **R^2^ (mean±SD) [95% CI]** | **rMSE (mean±SD) [95% CI]** |
| GP2 baseline | 0.23±0.002 [0.21, 0.25] | 0.04±0.001 [0.03, 0.05] | 1.06±0.011 [1.04, 1.08] |
| GP2 follow-up | 0.18±0.002 [0.16, 0.21] | 0.03±0.003 [0.02, 0.04] | 1.04±0.011 [1.02, 1.06] |
| CD | 0.35±0.013 [0.32, 0.39] | 0.10±0.008 [0.09, 0.13] | 1.22±0.55 [1.17, 1.23] |
| mean: mean scores across test sets; SD: standard deviation across test sets; *GP: general population subsamples; CD: clinical depression subsamples; CI: conference interval*. | | | |

**Supplemental Table 13**

| **Model performance in other profiles of sleep and depression** | | | |
| --- | --- | --- | --- |
| **Profiles** | **r (mean±SD) [95% CI]** | **R^2^ (mean±SD) [95% CI]** | **rMSE (mean±SD) [95% CI]** |
| DRS | 0.18±0.008 [0.17, 0.19] | 0.03±0.003 [0.03, 0.04] | 1.03±0.011 [1.03, 1.04] |
| NDRS | 0.16±0.011 [0.16, 0.17] | 0.03±0.004 [0.03, 0.03] | 0.99±0.009 [0.98, 0.99] |
| NSRD | 0.18±0.016 [0.17, 0.19] | 0.03±0.006 [0.03, 0.03] | 0.98±0.010 [0.98, 0.98] |
| Overall SH | 0.25±0.010 [0.25, 0.26] | 0.06±0.005 [0.06, 0.07] | 0.97±0.009 [0.96, 0.97] |
| Overall depression | 0.27±0.035 [0.30, 0.32] | 0.07±0.019 [0.09, 0.10] | 0.96±0.014 [0.75, 0.75] |
| mean: mean scores across test sets; *SD: standard deviation across test sets; DRS: depression-related SH, NDRS: non-depression-related SH, NSRD: non-SH-related depression, SH: sleep health; CI: conference interval.* | | | |

**Supplemental Table 14**

| **RCCA model performance in CD** | |
| --- | --- |
| **Subsamples** | **r (mean ± SD)** |
| Non-recurrent depression | 0.47 ± 0.006 |
| Untreated depression | 0.43 ± 0.011 |
| mean: mean scores across test sets; *CD: clinical depression subsamples; rCCA: regularized canonical correlation analysis* | |

**Supplemental Table 15**

| **Predictive model performance in CD** | | | |
| --- | --- | --- | --- |
| Subsamples | r (mean±SD) [95% CI] | R^2^ (mean±SD) [95% CI] | rMSE (mean±SD) [95% CI] |
| Non-recurrent depression (prediction) | 0.35±0.015 [0.32, 0.39] | 0.10±0.016 [0.09, 0.13] | 1.23±0.134 [1.18, 1.24] |
| Untreated depression (prediction) | 0.32±0.013 [0.29, 0.38] | 0.09±0.019 [0.07, 0.12] | 1.25±0.083 [1.19, 1.26] |
| mean: mean scores across test sets; *CD: clinical depression subsamples.* | | | |
